# Clear Cell Renal Cell Carcinoma: Unveiling Age-Linked Genetic Signatures, Relative Disease Aggressiveness, and Predictive Impact on Systemic Therapy

**DOI:** 10.1101/2024.12.30.24317177

**Authors:** Tanguy Pace-Loscos, Maeva Dufies, Renaud Schiappa, Manon Teisseire, Emmanuel Chamorey, Lisa Kinget, Benoit Beuselinck, Delphine Borchiellini, Gilles Pagès

## Abstract

**Background and objectives:** Age influences the development and progression of clear cell renal cell carcinoma (ccRCC). Management of non-metastatic (M0) ccRCC involves surgery, adjuvant immune checkpoint inhibition (e.g., pembrolizumab) and surveillance. For metastatic (M1) ccRCC, treatment combines immune checkpoint inhibitors with anti-angiogenic therapies, achieving long-term remission in approximately 30% of patients. Despite these advancements, a definitive cure remains elusive for most individuals. Furthermore, predictive criteria for relapse that incorporate age-specific variations are lacking, highlighting the critical need for prognostic models to optimize outcomes across diverse age groups. This study aimed to validate tumor aggressiveness markers in ccRCC, focusing on age as a determinant factor.

**Methods:** Data from The Cancer Genome Atlas (TCGA) were analyzed to identify age-related gene expression differences, which were then validated using an independent cohort and ccRCC cell lines. Random Survival Forest models were employed to predict overall survival (OS) and disease-free survival (DFS) post nephrectomy and progression-free survival upon tyrosine kinase inhibitors and immunotherapies.

**Key findings and limitations:** Of 110 age-differentiated genes identified, 31 were significantly correlated with OS and DFS. Poor prognostic markers included CIAO1, EIF2B1, NARF, UBE2G2, and SART3, while TMEM167B emerged as a good prognostic marker, and a potential predictive biomarker for immunotherapy efficacy. Limitations include the need for larger independent cohort validation and deeper exploration of gene mechanisms.

**Conclusions and clinical implications:** The study presents age-informed prognostic models that integrate gene expression profiles, offering a foundation for personalized treatment strategies. TMEM167B stands out as a novel candidate for improving age-specific ccRCC management.

**Advancing Practice:** **What does the study add?** This study explores age-dependent differentially expressed genes in clear cell renal cell carcinoma (ccRCC) and proposes a potential age-specific prognostic gene signature that could be applicable to both non-metastatic and metastatic patients. This signature may assist in identifying individuals at higher risk of relapse. Among the identified genes, *TMEM167B* is highlighted as a candidate associated with favorable prognosis and shows promise as a potential predictive biomarker for immunotherapy efficacy.

**Patient Summary:** Aggressiveness in ccRCC varies with age and gene expression. Tailored treatments based on these factors may enhance outcomes and minimize side effects, particularly for patients over 75 years of age.

## 1. Introduction

As global life expectancy increases, the prevalence of patients over 75 years of age with clear cell renal cell carcinoma (ccRCC) is also rising. Managing ccRCC in older patients with comorbidities presents challenges due to higher risks of adverse events associated with systemic and local treatments [1]. Although over half of ccRCC cases occur in individuals aged 65 and older, the specific characteristics of ccRCC in this population remain underexplored, complicating clinical decision-making. In younger patients, particularly those under 50, the behavior of ccRCC is a topic of ongoing debate. Some studies report comparable outcomes between younger and older patients[2, 3], while others suggest either more favorable [4, 5] or poorer outcomes [6] in younger populations. This age-related variability underscores the importance of personalized therapeutic approaches that consider both tumor biology and patient age [7, 8].

In non-metastatic (M0) ccRCC, treatment relies on surgery then surveillance or adjuvant immune checkpoint inhibitors (ICIs) [9]. However, questions remain about stage-specific treatment strategies and the stratification of patients based on age and biomarkers.

In metastatic (M1) ccRCC, anti-angiogenic therapies have improved progression-free survival (PFS) [10] and combining ICIs with anti-angiogenic agents [11] has further improved overall survival (OS). Dual ICI combinations have also demonstrated notable improvements in OS [12]. However, these approaches require monitoring in elderly patients to mitigate toxicity.

This study investigates whether age impacts disease-free survival (DFS), PFS, and OS post-nephrectomy or post-start of systemic therapy in ccRCC patients and identifies genetic markers to stratify patients by risk and optimize therapies. Tailored therapeutic strategies are essential for managing ccRCC in young and elderly patients. Understanding age-related genetic and molecular differences in ccRCC could lead to personalized treatments, improving outcomes and reducing adverse effects. Defining genetic markers for ccRCC aggressiveness and therapy response is critical for advancing precision medicine in frail, elderly populations.

## 2. Patients and methods

### 2.1. Datasets sources and processing

RNA sequencing data from 537 ccRCC patients (449 M0 and 84 M1 patients) were obtained from The Cancer Genome Atlas (TCGA) KIRC dataset (https://portal.gdc.cancer.gov/). The patient population was randomly divided into training and validation cohorts.

Cohort of metastatic patients (M1) either treated with anti-angiogenic drugs or ICIs were described previously [13]. We utilized the GEPIA (Gene Expression Profiling Interactive Analysis) software http://gepia2.cancer-pku.cn/#index to compare the expression levels of specific genes in normal tissues versus tumor tissues.

### 2.2. Variables definitions

- Age and raw counts of RNA-sequencing data were available for analysis. Patients were categorized into four distinct age groups:
- Patients < 50 years old
- Patients between 50 and 65 years old
- Patients between 65 and 75 years old
- Patients ≥ 75 years old

### 2.3. Genes selection

The selection of genes was based on comparing the first (Q1) and third (Q3) quartiles of RNA-seq counts for the entire population to the median counts of each of the four age groups. A gene was selected if the median count for any subgroup met either of the following criteria:

- The subgroup median was less than 110% of the overall Q1.
- The subgroup median was greater than 95% of the overall Q3.

### 2.4. Outcomes

The study investigated the predictive score for OS, DFS and PFS based on gene expression.

- **OS**: For M0 patients, OS at 60 months was defined as the time from surgery to death from any cause. For M1 patients, OS at 24 months was defined as the time from diagnosis to death.
- **DFS**: For M0 patients, DFS at 36 months was defined as the time from surgery to the occurrence of metastasis. For M1 patients, DFS at 12 months was defined as the time from diagnosis to relapse.
- **PFS**: For M1 patients, PFS at 12 months was defined as the duration from the initiation of systemic treatment to relapse.

### 2.5. Statistical analysis

Categorical data were presented as frequencies, while continuous data included mean, median, minimum, maximum, and standard deviation. Principal Component Analysis (PCA) assessed collinearity among variables. Statistical tests, including T-tests, Wilcoxon rank sum, ANOVA, or Kruskal-Wallis, assessed variable distributions and relationships, while missing data were reported as absolute numbers and percentages. Censored data were presented with the median follow-up, calculated using the inverse Kaplan-Meier method, and illustrated with a Kaplan-Meier curve [14]. Survival percentages with 95% confidence intervals were calculated at specified intervals. Survival curves were compared using the Log-Rank test, and hazard ratios were estimated via Cox regression.

Prognostic variables for OS, DFS, and PFS were identified using univariate Cox regression, with FDR-adjusted P-values (<0.05) included in a random survival forest (RSF) model. The final model generated a classifier to predict the risk of events. The optimal classifier was determined by selecting the linear predictor that maximized the sum of sensitivity and specificity, with a minimum sensitivity of 80%. The resulting threshold was used to distinguish between low- and high-risk patient groups. Variables identified from the RSF model were then incorporated into a multivariate Cox regression analysis to calculate hazard ratios (HR) and 95% confidence intervals (CI). The best multivariate Cox model was selected using a stepwise algorithm guided by Akaike’s Information Criterion (AIC) [15]. All statistical analyses were conducted using R version 4.3.1. The *rfsrc* function from the random Forest SRC library was employed to build the random forest survival model. Bestcut2 function from greyzoneSurv library used to find cut-off for continuous variables in bivariate survival analysis. Our results adhere to the REMARK reporting criteria [16].

### 2.6. Cell lines

Human ccRCC cell lines, 786-O, A498, and ACHN were purchased from the American Tissue Culture Collection (ATCC) and RCC10 were a kind gift from W.H. Kaelin (Dana-Farber Cancer Institute, Boston, MA). Human Renal Epithelial Cells (HREPC) were purchased from Promocell (Heidelberg Germany). Primary human ccRCC cells were isolated by enzymatic dissociation from surgical specimens (CHU Nice, Department of Pathology) and cultured in PromoCell Renal Epithelial Cell Growth Medium 2. Analysis of mRNA levels by quantitative real-time PCR (qPCR) experiments was performed as previously described [17].

## 3. Results

### 3.1. Differential Gene Expression Across Age Groups in the M0 Population

To reduce confusion from factors unique to either M0 or M1 disease, this study focuses first on the M0 population (449 patients), analyzing differential gene expression across defined age groups. For those patients mean age was 60.7 years, median follow-up from diagnosis was 50.1 months (CI95% [48.8;57.6]), median survival was not reached. Most patients were male (63.7%), T1 (50%), N0 (94.8). Tumor with Fuhrman grade 2 and 3 were preponderant (respectively 49.7% and 39%, Supplementary Table 1). Among 30,000 genes, 110 were differentially expressed in the four distinct age groups (Table 1).

**Table 1:**
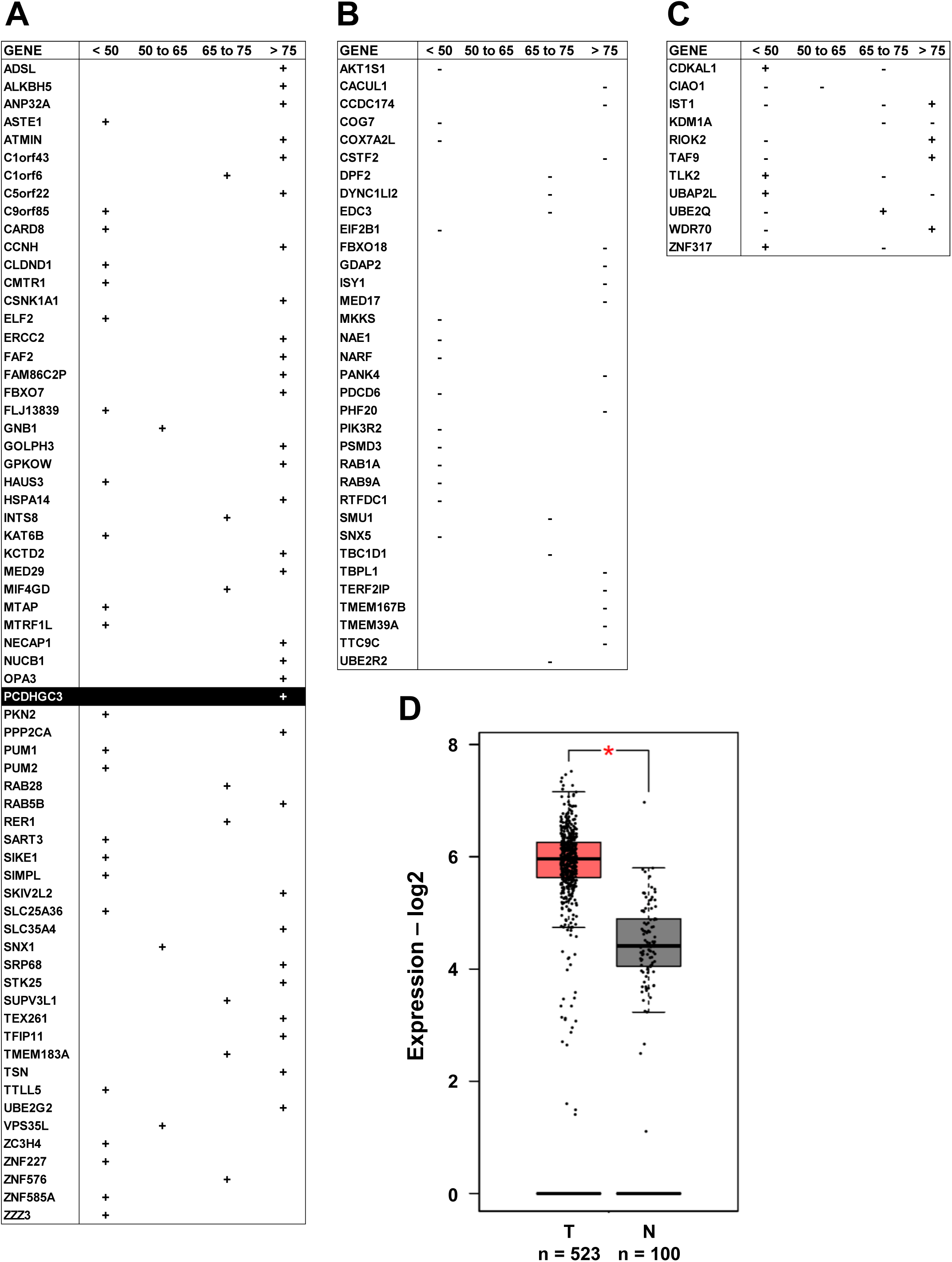
Differentially Expressed Genes in the General Genome. A “–” sign indicates decreased expression in a specific tranche (A), while a “+” sign denotes increased expression (B). Certain genes exhibit variable expressions, showing either an increase or decrease depending on the specific tranche (C). D) Relative expression of PCDHGC3 in norma and tumor tissue according to the GEPIA software (* P < 0.05).

| GENE | < 50 | 50 to 65 | 65 to 75 | > 75 |
| --- | --- | --- | --- | --- |
| ADSL |  |  |  | + |
| ALKBH5 |  |  |  | + |
| ANP32A |  |  |  | + |
| ASTE1 | + |  |  |  |
| ATMIN |  |  |  | + |
| C1orf43 |  |  |  | + |
| C1orf6 |  |  | + |  |
| C5orf22 |  |  |  | + |
| C9orf85 | + |  |  |  |
| CARD8 | + |  |  |  |
| CCNH |  |  |  | + |
| CLDND1 | + |  |  |  |
| CMTR1 | + |  |  |  |
| CSNK1A1 |  |  |  | + |
| ELF2 | + |  |  |  |
| ERCC2 |  |  |  | + |
| FAF2 |  |  |  | + |
| FAM86C2P |  |  |  | + |
| FBXO7 |  |  |  | + |
| FLJ13839 | + |  |  |  |
| GNB1 |  | + |  |  |
| GOLPH3 |  |  |  | + |
| GPKOW |  |  |  | + |
| HAUS3 | + |  |  |  |
| HSPA14 |  |  |  | + |
| INTS8 |  |  | + |  |
| KAT6B | + |  |  |  |
| KCTD2 |  |  |  | + |
| MED29 |  |  |  | + |
| MIF4GD |  |  | + |  |
| MTAP | + |  |  |  |
| MTRF1L | + |  |  |  |
| NECAP1 |  |  |  | + |
| NUCB1 |  |  |  | + |
| OPA3 |  |  |  | + |
| PCDHGC3 |  |  |  | + |
| PKN2 | + |  |  |  |
| PPP2CA |  |  |  | + |
| PUM1 | + |  |  |  |
| PUM2 | + |  |  |  |
| RAB28 |  |  | + |  |
| RAB5B |  |  |  | + |
| RER1 |  |  | + |  |
| SART3 | + |  |  |  |
| SIKE1 | + |  |  |  |
| SIMPL | + |  |  |  |
| SKIV2L2 |  |  |  | + |
| SLC25A36 | + |  |  |  |
| SLC35A4 |  |  |  | + |
| SNX1 |  | + |  |  |
| SRP68 |  |  |  | + |
| STK25 |  |  |  | + |
| SUPV3L1 |  |  | + |  |
| TEX261 |  |  |  | + |
| TFIP11 |  |  |  | + |
| TMEM183A |  |  | + |  |
| TSN |  |  |  | + |
| TTLL5 | + |  |  |  |
| UBE2G2 |  |  |  | + |
| VPS35L |  | + |  |  |
| ZC3H4 | + |  |  |  |
| ZNF227 | + |  |  |  |
| ZNF576 |  |  | + |  |
| ZNF585A | + |  |  |  |
| ZZZ3 | + |  |  |  |

B
| GENE | < 50 | 50 to 65 | 65 to 75 | > 75 |
| --- | --- | --- | --- | --- |
| AKT1S1 | - |  |  |  |
| CACUL1 |  |  |  | - |
| CCDC174 |  |  |  | - |
| COG7 | - |  |  |  |
| COX7A2L | - |  |  |  |
| CSTF2 |  |  |  | - |
| DPF2 |  |  | - |  |
| DYNC1LI2 |  |  | - |  |
| EDC3 |  |  | - |  |
| EIF2B1 | - |  |  |  |
| FBXO18 |  |  |  | - |
| GDAP2 |  |  |  | - |
| ISY1 |  |  |  | - |
| MED17 |  |  |  | - |
| MKKS | - |  |  |  |
| NAE1 | - |  |  |  |
| NARF | - |  |  |  |
| PANK4 |  |  |  | - |
| PDCD6 | - |  |  |  |
| PHF20 |  |  |  | - |
| PIK3R2 | - |  |  |  |
| PSMD3 | - |  |  |  |
| RAB1A | - |  |  |  |
| RAB9A | - |  |  |  |
| RTFDC1 | - |  |  |  |
| SMU1 |  |  | - |  |
| SNX5 | - |  |  |  |
| TBC1D1 |  |  | - |  |
| TBPL1 |  |  |  | - |
| TERF2IP |  |  |  | - |
| TMEM167B |  |  |  | - |
| TMEM39A |  |  |  | - |
| TTC9C |  |  |  | - |
| UBE2R2 |  |  | - |  |

C
| GENE | < 50 | 50 to 65 | 65 to 75 | > 75 |
| --- | --- | --- | --- | --- |
| CDKAL1 | + |  | - |  |
| CIAO1 | - | - |  |  |
| IST1 | - |  | - | + |
| KDM1A |  |  | - | - |
| RIOK2 | - |  |  | + |
| TAF9 | - |  |  | + |
| TLK2 | + |  | - |  |
| UBAP2L | + |  |  | - |
| UBE2Q | - |  | + |  |
| WDR70 | - |  |  | + |
| ZNF317 | + |  | - |  |

In individuals under 50, more genes were up-regulated (e.g., ASTE1, ELF2, KAT6B, ZZZ3) than down-regulated (e.g., AKT1S1, EIF2B1, NARF). Between 50 and 65 years old (Table 1A-B), only GNB1, SNX1, and VPS35L were up-regulated, with no down-regulation. For ages 65–75, eight genes were up-regulated (e.g., INTS8, TMEM183A) and six were down-regulated (e.g., DPF2, TBC1D1). In those over 75 (Table 1C), a distinct pattern emerged with genes such as ADSL, FBXO7, and PPP2CA up-regulated, while others like CSTF2 and TMEM167B were down-regulated. Some genes exhibited inverse regulation across age groups (e.g., CDKAL1, TLK2, ZNF317), indicating age-related shifts in cellular processes.

IST1 was down-regulated in individuals under 50 and 65–75, yet up-regulated in those over 75. Protocadherin gamma-C3 (PCDHGC3) was elevated in tumor tissues, suggesting a role in tumor progression (Table 1D). TMEM167B, linked to the Golgi apparatus, showed no prior functional studies, marking this as a novel finding.

These age-stratified expression patterns suggest that aging influences gene expression dynamics in ccRCC. Younger individuals showed broader up-regulation of genes, while older groups exhibited distinct shifts in regulation, indicating possible changes in cellular pathways. Genes like RIOK2, TAF9, and WDR70 showed opposite trends in younger versus older individuals, highlighting their potential role in aging-related biological mechanisms. This is the first comprehensive panel identifying differentially expressed genes across age groups in ccRCC, offering insights into age-associated gene expression and its implications for disease progression and therapeutic targeting.

### 3.2. Correlation Between Identified Genes and Survival Outcomes

We investigated the relationship between gene expression and survival outcomes, focusing on OS and DFS. Overexpression of six genes genes-EIF2B1, NARF, INTS8, C1orf6, PCDHGC3 and UBE2Q-was associated with shorter OS and DFS, identifying them as markers of tumor aggressiveness and poor prognosis. Conversely, genes such as SMU1, FBXO7, MTAP and C9orf85 were correlated with longer OS and DFS suggesting lower tumor aggressiveness and a more favorable prognosis (Table 2 and Table 3). Age-related variations in gene expression were also significant, emphasizing their role in survival outcomes and their potential as therapeutic targets for ccRCC management (Supplementary Fig. 1 and Supplementary Fig. 2).

**Table 2:**
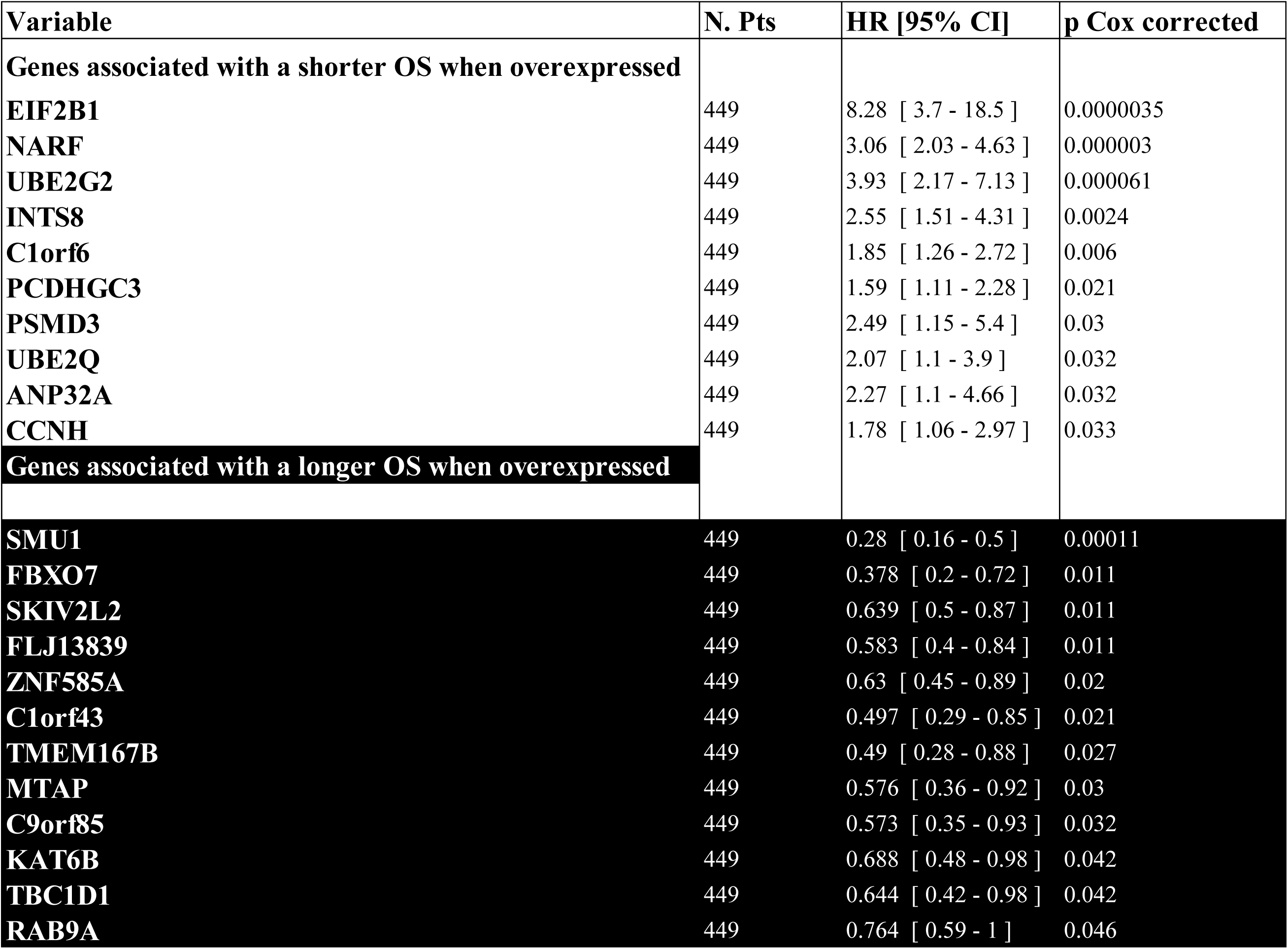
Overall survival rates for the different genes. HR values with 95% CI are indicated along with P values. Genes associated with a poor prognosis when overexpressed are indicated on a white font whereas genes associated with a longer OS are indicated on a black font.

| Variable | N. Pts | HR [95% CI] | p Cox corrected |
| --- | --- | --- | --- |
| **Genes associated with a shorter OS when overexpressed** |  |  |  |
| **EIF2B1** | 449 | 8.28 [ 3.7 - 18.5 ] | 0.0000035 |
| **NARF** | 449 | 3.06 [ 2.03 - 4.63 ] | 0.000003 |
| **UBE2G2** | 449 | 3.93 [ 2.17 - 7.13 ] | 0.000061 |
| **INTS8** | 449 | 2.55 [ 1.51 - 4.31 ] | 0.0024 |
| **C1orf6** | 449 | 1.85 [ 1.26 - 2.72 ] | 0.006 |
| **PCDHGC3** | 449 | 1.59 [ 1.11 - 2.28 ] | 0.021 |
| **PSMD3** | 449 | 2.49 [ 1.15 - 5.4 ] | 0.03 |
| **UBE2Q** | 449 | 2.07 [ 1.1 - 3.9 ] | 0.032 |
| **ANP32A** | 449 | 2.27 [ 1.1 - 4.66 ] | 0.032 |
| **CCNH** | 449 | 1.78 [ 1.06 - 2.97 ] | 0.033 |
| **Genes associated with a longer OS when overexpressed** |  |  |  |
| **SMU1** | 449 | 0.28 [ 0.16 - 0.5 ] | 0.00011 |
| **FBXO7** | 449 | 0.378 [ 0.2 - 0.72 ] | 0.011 |
| **SKIV2L2** | 449 | 0.639 [ 0.5 - 0.87 ] | 0.011 |
| **FLJ13839** | 449 | 0.583 [ 0.4 - 0.84 ] | 0.011 |
| **ZNF585A** | 449 | 0.63 [ 0.45 - 0.89 ] | 0.02 |
| **C1orf43** | 449 | 0.497 [ 0.29 - 0.85 ] | 0.021 |
| **TMEM167B** | 449 | 0.49 [ 0.28 - 0.88 ] | 0.027 |
| **MTAP** | 449 | 0.576 [ 0.36 - 0.92 ] | 0.03 |
| **C9orf85** | 449 | 0.573 [ 0.35 - 0.93 ] | 0.032 |
| **KAT6B** | 449 | 0.688 [ 0.48 - 0.98 ] | 0.042 |
| **TBC1D1** | 449 | 0.644 [ 0.42 - 0.98 ] | 0.042 |
| **RAB9A** | 449 | 0.764 [ 0.59 - 1 ] | 0.046 |

**Table 3:** Disease-free survival for the different genes. HR values with 95% CI are indicated along with P values. Genes associated with a poor prognosis when overexpressed are indicated on a white font, whereas genes associated with a longer DFS are indicated on a black font.

| Variable | N. Pts | HR [95% CI] | p Cox corrected |
| --- | --- | --- | --- |
| **Genes associated with a shorter DFS when overexpressed** |  |  |  |
| **NARF** | 379 | 2.75 [ 1.53 - 4.94 ] | 0.0032 |
| **EIF2B1** | 379 | 5.66 [ 1.99 - 16.1 ] | 0.0041 |
| **C1orf6** | 379 | 2.08 [ 1.27 - 3.39 ] | 0.0092 |
| **AKT1S1** | 379 | 2.3 [ 1.3 - 4.05 ] | 0.0092 |
| **PCDHGC3** | 379 | 2.09 [ 1.26 - 3.47 ] | 0.0092 |
| **CSNK1A1** | 379 | 2.83 [ 1.38 - 5.79 ] | 0.0092 |
| **CIAO1** | 379 | 4.8 [ 1.61 - 14.3 ] | 0.0092 |
| **INTS8** | 379 | 2.7 [ 1.34 - 5.42 ] | 0.0092 |
| **UBE2Q** | 379 | 2.77 [ 1.21 - 6.34 ] | 0.025 |
| **NUCB1** | 379 | 2.41 [ 1.14 - 5.1 ] | 0.028 |
| **UBAP2L** | 379 | 3.1 [ 1.19 - 8 ] | 0.028 |
| **SART3** | 379 | 2.97 [ 1.12 - 7.9 ] | 0.036 |
| **Genes associated with a longer DFS when overexpressed** |  |  |  |
| **SMU1** | 379 | 0.17 [ 0.076 - 0.36 ] | 0.00012 |
| **C9orf85** | 379 | 0.34 [ 0.18 - 0.63 ] | 0.0032 |
| **FBXO7** | 379 | 0.22 [ 0.09 - 0.52 ] | 0.0032 |
| **MTAP** | 379 | 0.36 [ 0.2 - 0.63 ] | 0.0032 |
| **UBE2R2** | 379 | 0.31 [ 0.12 - 0.81 ] | 0.025 |
| **SIMPL** | 379 | 0.77 [ 0.6 - 1 ] | 0.046 |
| **MTRF1L** | 379 | 0.49 [ 0.25 - 1 ] | 0.046 |
| **OPA3** | 379 | 0.57 [ 0.33 - 0.99 ] | 0.046 |
| **TBC1D1** | 379 | 0.55 [ 0.31 - 1 ] | 0.049 |

### 3.3. Data quality

Principal component analysis (PCA) of selected genes for OS and DFS revealed variance spread across 10 dimensions, without distinct patient groups emerging (Supplementary Fig. 3). The first dimension contributed slightly more for OS but was insufficient for classification (Supplementary Fig. 3A-C). Minimal outliers indicated robust data (Supplementary Fig. 3B-D), underscoring the complexity of gene expression in ccRCC survival outcomes.

### 3.4. Univariate analyses of clinical parameters for OS and DFS

Univariate analyses were conducted to evaluate the relationship between clinical parameters, including sex, Fuhrman grade (combining grades 1 and 2 vs. grades 3 and 4), tumor size (T1 + T2 vs. T3), and tumor stage. Stage III, T3, and Fuhrman grade 3 were associated with shorter survival, while sex showed no significant correlation (Supplementary Table 2).

Equivalent analyses were conducted for DFS. Tumor stages II and III and T3 were associated with shorter DFS, while sex and grade showed no correlation (Supplementary Table 3).

### 3.5. Prognostic score for OS; M0 patients

To develop a prognostic score for OS, multivariate analyses with selected genes (Table 2) and clinical parameters were performed using the RSF method. The model had an optimal threshold of 8.156251, with 98% sensitivity and 79% specificity in the training cohort, and 67% sensitivity and 65% specificity in the validation cohort (Supplementary Fig. 4). High-risk patients had shorter OS at 60 months (HR: 3.4, 95% CI: [1.5–7.5], P = 0.00231), with survival at 56% compared to 83% in low-risk patients (Fig. 1A, and Supplementary Fig. 4). Tumor stage, age, NARF, UBE2G2, INTS8, ANP32 and TBC1D1 predicted poor outcomes, while TMEM167B indicated better prognosis (Supplementary Table 4).

**Fig. 1:**
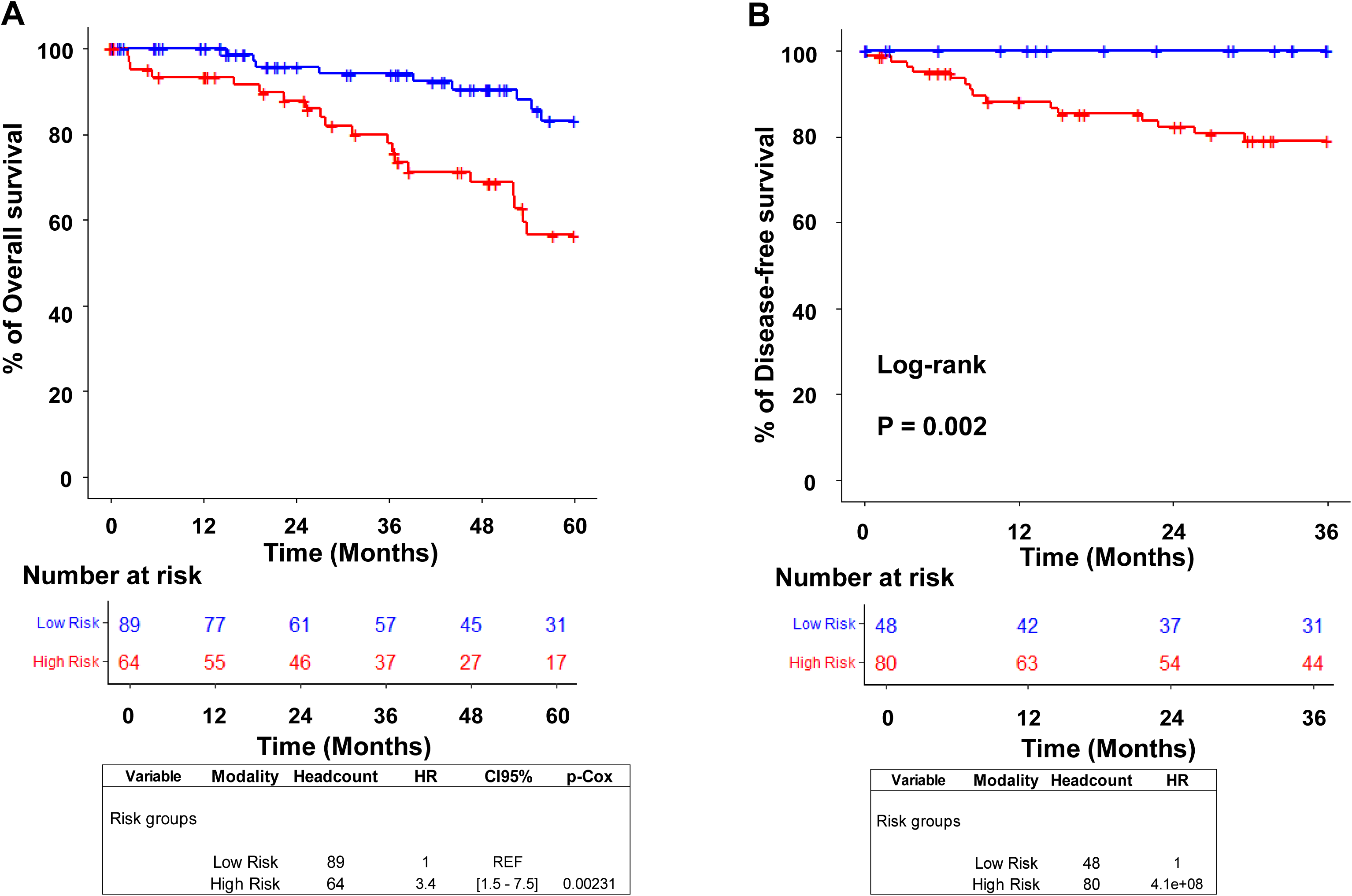
OS and DFS Curves for Low- and High-Risk M0 Patients. A) Kaplan–Meier analysis of OS in low-risk and high-risk patient groups. The number of patients in each group is indicated, along with the corresponding P-value. B) Kaplan–Meier analysis of DFS in low-risk and high-risk patient groups. The number of patients in each group is indicated, along with the corresponding P-value. As all patients are assigned to the right risk group HR and cox P-value are not calculable. LogRank test was performed in place of Cox bivariate regression.

### 3.6. Prognostic score for DFS; M0 patients

The DFS prognostic score was developed as for the OS prognostic score with selected genes (Table 3) and clinical parameters via the RSF method. The training cohort achieved 100% sensitivity and 86% specificity, while the validation cohort maintained 100% sensitivity but had 42% specificity (Supplementary Fig. 5). Patients were stratified into low- and high-risk groups, with high-risk patients showing a DFS rate of 79% compared to 100% for low-risk patients (Fig. 1B and Supplementary Table 5). Cox regression linked tumor stage, C1orf6, CIAO1, and INTS8 to poor prognosis, while MTAP indicated better outcomes.

The functions of genes contributing to the prognostic scores for OS and DFS are detailed in the supplementary materials, showcasing their diverse roles and providing novel insights into previously uncharacterized genes such as TMEM167B.

### 3.7. Power of the genes of interest and of the model depending on age tranches for OS and DFS: M0 patients

Our model demonstrated associations with OS across all age groups, with the strongest relevance observed in patients over 50 years old (P < 0.01), particularly those aged 50-65 and 65-75 years (P < 0.001). For DFS, the model showed associations across all age tranches, especially in patients aged above 50 (P < 0.0001). These findings highlight the model’s predictive strength, particularly for older populations, (Supplementary Fig. 6).

### 3.8. Prognostic score for OS and DFS in M1 patients

The prognostic model developed for M0 patients was tested on M1 patients (Supplementary Table 6). PCA revealed no distinct M1 patient groups (Supplementary Fig. 8). Using the RSF method, the model demonstrated high specificity and sensitivity for both OS and DFS (Supplementary Fig. 9 and Supplementary Fig. 10). High-risk M1 patients had shorter OS (HR: 9.2, P < 0.0001) with a 2% survival rate at 24 months (Fig. 2A), and shorter DFS (Fig. 2B) with a 6% rate at 12 months. This model effectively identifies high-risk M1 patients.

**Fig. 2:**
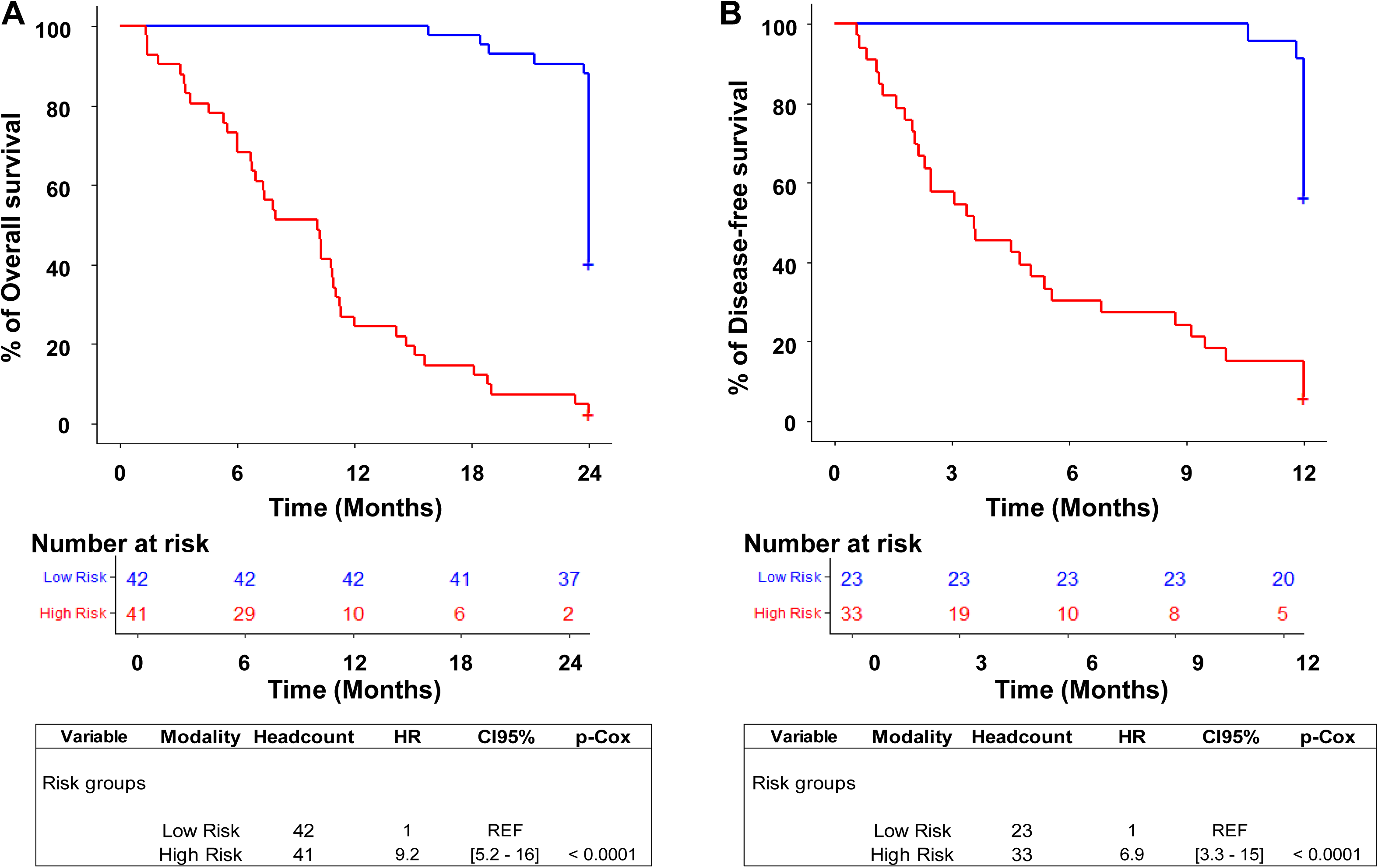
OS and DFS Curves for Low- and High-Risk M1 Patients. A) Kaplan–Meier analysis of OS in low-risk and high-risk patient groups. The number of patients in each group is indicated, along with the corresponding P-value. B) Kaplan–Meier analysis of DFS in low-risk and high-risk patient groups. The number of patients in each group is indicated, along with the corresponding P-value.

### 3.9. Power of the model depending on age tranches for OS and DFS for M1 patients

The relevance of our model was assessed by analyzing its association with OS across different age groups. The model demonstrated a significant correlation with OS in aged under 50 years (P = 0.00051), aged 50-65 (P < 0.001) and 65-75 years (P < 0.0001). The significant P values suggest that the model is most relevant for the population aged less than 75 years old (Supplementary Fig. 11).

The same model was applied to DFS. The P values appeared significant for patients aged 50-60 years (P < 0.0001), 60-75 years (P < 0.0001), and over 75 years (P = 0.0066). The P values < 0.0001 suggests that the model is most relevant for the population aged above 50 years old (Supplementary Fig. 12). The sample size of patients aged above 75 years was insufficient to conduct robust statistical analysis.

### 3.10. TMEM167B as a Promising Gene for Future Exploratory Studies

Among 110 differentially expressed genes, TMEM167B lacked prior research, prompting further studies. Its mRNA levels were lower in ccRCC cells than in normal renal epithelial cells, aligning with a favorable prognosis (Fig. 3A). While TMEM167B showed no significant impact on OS or PFS in patients treated with anti-angiogenic TKIs (Fig. 3B), high expression correlated with longer OS and PFS in patients receiving ICIs (Fig. 3C). These findings highlight TMEM167B as both a prognostic marker and a predictor of immunotherapy efficacy.

**Fig. 3:**
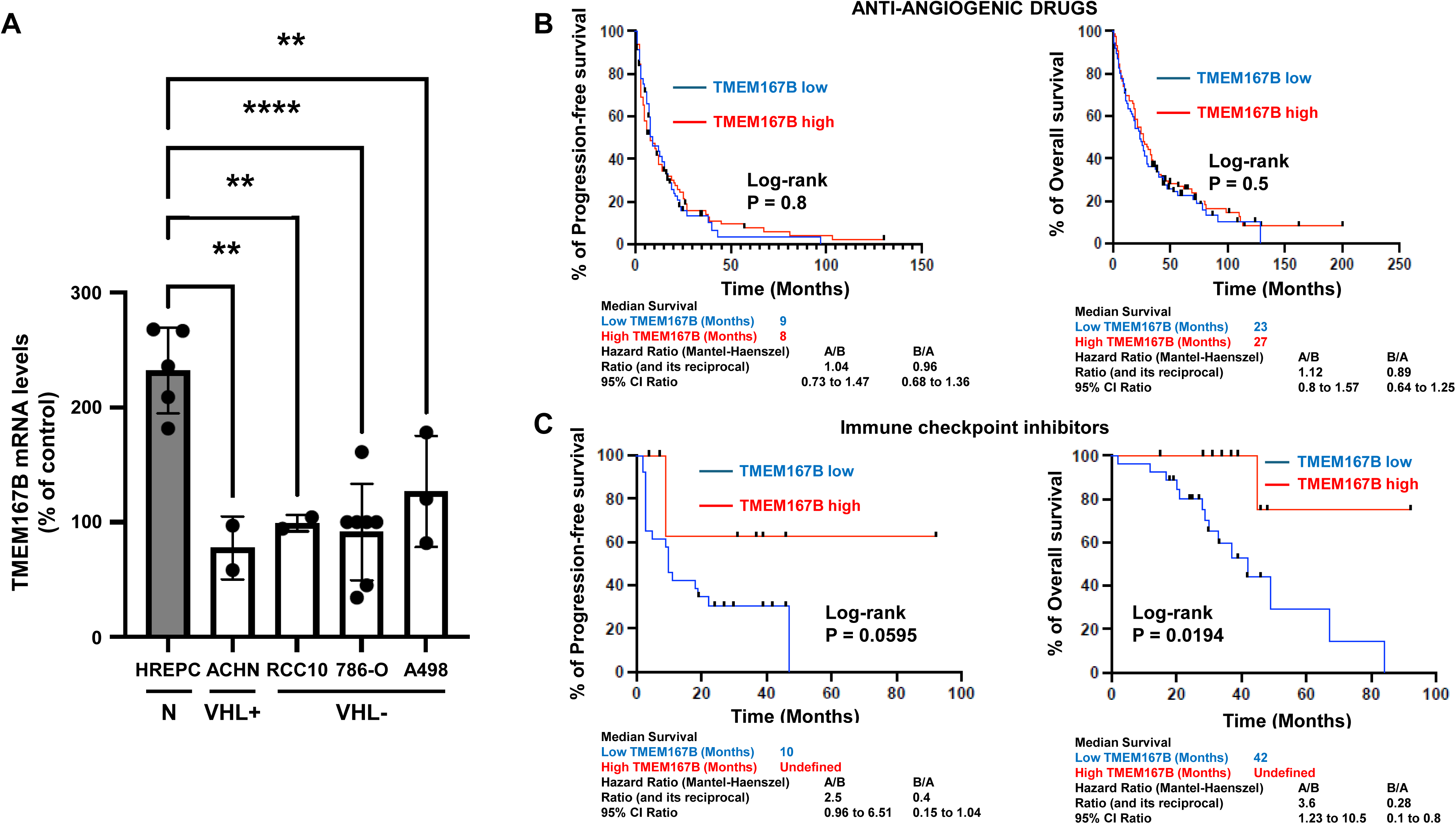
TMEM167B is expressed to a lower extent in ccRCC cell lines and is predictive of efficacy of immune checkpoint inhibitors. A) mRNA levels were assed in normal epithelial cells (HREPC) and in ccRCC cell line that were either wild-type or mutated for the VHL gene. B) Relationship to progression-free survival and overall survival for patient treated by anti-angiogenic drugs. P value HR and 95% CI are indicated. C) Relationship to progression-free survival and overall survival for patients treated by immune checkpoint inhibitors. P value HR and 95% CI are indicated.

## 4. Discussion

Despite advances in ccRCC treatment, challenges remain, especially for older patients, who form most diagnoses between ages 65 and 70. This demographic is often overlooked in discussions about personalized therapy, despite the growing need to balance treatment efficacy with quality of life in an aging population. Tailoring treatments to patient frailty, often age-dependent, is crucial. Our study addresses these concerns by analyzing publicly available data, identifying age-associated differential gene expressions among 30,000 genes. Notably, many age-related genes identified are recognized as good prognostic factors, highlighting the potential for age-specific treatment strategies to improve outcomes and meet the unique needs of older ccRCC patients.

### Development of an Age-Specific Gene Signature

We developed a novel age-specific gene signature with relevance to OS and DFS in diverse M0 patients, validated in TCGA data. It showed the strongest OS significance in patients over 50, while DFS significance peaked in those aged 50–75. The signature also proved effective for M1 patients in the same age group. These findings highlight its potential for identifying high-risk patients who may benefit from tailored monitoring and treatments. However, further validation is essential before clinical implementation.

### The Role of TMEM167B in Personalized Therapy

TMEM167B stands out as a promising predictive marker for immunotherapy efficacy. Its role in ICIs resistance remains underexplored. This discovery opens new research avenues to better understand and overcome resistance to immunotherapy.

### Reversal of Hazard Ratio for TBC1D1

The HR for high expression of TBC1D1 shifted from a protective effect in the univariate analysis to a risk-enhancing effect in the multivariate Cox model. This change is likely attributable to the high collinearity inherent in gene expression data. Adjusting for other covariates in the multivariate model may have uncovered interactions or confounding effects, leading to the observed reversal in the relationship. This phenomenon is consistent with previous studies emphasizing the substantial collinearity in genomic data, which can obscure or even invert associations in survival analyses when accounting for multiple predictors [18].

## 5. Conclusions

This study proposes a methodology to improve ccRCC treatment by emphasizing the role of age in addressing older patients’ therapeutic needs. It highlights the need for further research to validate findings, explore *TMEM167B*, and refine age-specific strategies. The next step involves comparing the model’s predictive power with established nomograms (Leibovich score [19], MSKCC and IMDC scores [20]), from the European Association of Urology, which use only clinical and histological parameters. This work lays the foundation for personalized, age-adapted therapeutic approaches.

## Author contributions

Gilles Pagès and Tanguy Pacé-Loscos had full access to all the data in the study and took responsibility for the integrity of the data and the accuracy of the data analysis.

***Study concept and design***: Gilles Pagès.

***Acquisition of data***: Renaud Schiappa, Benoit Beuselinck, Lisa Kinget.

***Analysis and interpretation of data***: Tanguy Pacé-Loscos, Maeva Dufies and Gilles Pagès.

***Drafting of the manuscript***: Gilles Pagès and Tanguy Pacé-Loscos.

***Critical revision of the manuscript for important intellectual content***: Tanguy Pacé-Loscos, Maeva Dufies, Renaud Schiappa, Manon Teisseire, Emmanuel Chamorey, Gilles Pagès, Delphine Borchiellini, Benoit Beuselinck, Lisa Kinget.

***Statistical analysis***: Tanguy Pacé-Loscos,

***Obtaining funding***: Gilles Pagès

***Administrative, technical, or material support***: None.

***Supervision***: Gilles Pagès.

## Data Availability

All data produced in the present work are contained in the manuscript

https://www.cbioportal.org/

## References

[1] Iqbal M. Renal Cell Carcinoma: A Complex Therapeutic Challenge in the Elderly. Cureus. 2022;14:e26346.

[2] Hermansen CK, Donskov F. Outcomes based on age in patients with metastatic renal cell carcinoma treated with first line targeted therapy or checkpoint immunotherapy: Older patients more prone to toxicity. J Geriatr Oncol. 2021;12:827–33.

[3] Thompson RH, Ordonez MA, Iasonos A, Secin FP, Guillonneau B, Russo P, et al. Renal cell carcinoma in young and old patients--is there a difference? J Urol. 2008;180:1262–6; discussion 6.

[4] Taccoen X, Valeri A, Descotes JL, Morin V, Stindel E, Doucet L, et al. Renal cell carcinoma in adults 40 years old or less: young age is an independent prognostic factor for cancer-specific survival. Eur Urol. 2007;51:980–7.

[5] Sanchez-Ortiz RF, Rosser CJ, Madsen LT, Swanson DA, Wood CG. Young age is an independent prognostic factor for survival of sporadic renal cell carcinoma. J Urol. 2004;171:2160–5.

[6] Zhang G, Zhu Y, Dong D, Gu W, Zhang H, Sun L, et al. Clinical outcome of advanced and metastatic renal cell carcinoma treated with targeted therapy: is there a difference between young and old patients? Onco Targets Ther. 2014;7:2043–52.

[7] Tomita Y, Motzer RJ, Choueiri TK, Rini BI, Miyake H, Uemura H, et al. Efficacy and safety of avelumab plus axitinib in elderly patients with advanced renal cell carcinoma: extended follow-up results from JAVELIN Renal 101. ESMO Open. 2022;7:100450.

[8] Kirkali Z. Kidney cancer in the elderly. Urol Oncol. 2009;27:673–6.

[9] Choueiri TK, Tomczak P, Park SH, Venugopal B, Ferguson T, Chang YH, et al. Adjuvant Pembrolizumab after Nephrectomy in Renal-Cell Carcinoma. N Engl J Med. 2021;385:683–94.

[10] Haibe Y, Kreidieh M, El Hajj H, Khalifeh I, Mukherji D, Temraz S, et al. Resistance Mechanisms to Anti-angiogenic Therapies in Cancer. Front Oncol. 2020;10:221.

[11] Rini BI, Plimack ER, Stus V, Gafanov R, Hawkins R, Nosov D, et al. Pembrolizumab plus Axitinib versus Sunitinib for Advanced Renal-Cell Carcinoma. N Engl J Med. 2019;380:1116–27.

[12] Motzer RJ, Tannir NM, McDermott DF, Aren Frontera O, Melichar B, Choueiri TK, et al. Nivolumab plus Ipilimumab versus Sunitinib in Advanced Renal-Cell Carcinoma. N Engl J Med. 2018;378:1277–90.

[13] Kinget L, Naulaerts S, Govaerts J, Vanmeerbeek I, Sprooten J, Laureano RS, et al. A spatial architecture-embedding HLA signature to predict clinical response to immunotherapy in renal cell carcinoma. Nat Med. 2024;30:1667–79.

[14] Schemper M, Smith TL. A note on quantifying follow-up in studies of failure time. Control Clin Trials. 1996;17:343–6.

[15] Akaike H. Information Theory and an Extension of the Maximum Likelihood Principle. 1973.

[16] McShane LM, Altman DG, Sauerbrei W, Taube SE, Gion M, Clark GM, et al. REporting recommendations for tumour MARKer prognostic studies (REMARK). Br J Cancer. 2005;93:387–91.

[17] Dufies M, Giuliano S, Ambrosetti D, Claren A, Ndiaye PD, Mastri M, et al. Sunitinib Stimulates Expression of VEGFC by Tumor Cells and Promotes Lymphangiogenesis in Clear Cell Renal Cell Carcinomas. Cancer Res. 2017;77:1212–26.

[18] Mohammed M, Mboya IB, Mwambi H, Elbashir MK, Omolo B. Predictors of colorectal cancer survival using cox regression and random survival forests models based on gene expression data. PLoS One. 2021;16:e0261625.

[19] Leibovich BC, Blute ML, Cheville JC, Lohse CM, Frank I, Kwon ED, et al. Prediction of progression after radical nephrectomy for patients with clear cell renal cell carcinoma: a stratification tool for prospective clinical trials. Cancer. 2003;97:1663–71.

[20] Okita K, Hatakeyama S, Tanaka T, Ikehata Y, Tanaka T, Fujita N, et al. Impact of Disagreement Between Two Risk Group Models on Prognosis in Patients With Metastatic Renal-Cell Carcinoma. Clin Genitourin Cancer. 2019;17:e440–e6.

