## Supplementary Text Figures and Tables for "Clear Cell Renal Cell Carcinoma: Unveiling Age-Linked Genetic Signatures, Relative Disease Aggressiveness, and Predictive Impact on Systemic Therapy"

#### Supplemental materials

##### Function of the different genes of the signature

**The Proline-rich AKT1 substrate 1** (**AKT1S1**) gene encodes a proline-rich substrate of AKT. It regulates the activity of the mTORC1 complex [1, 2]. Its activity is dependent on its phosphorylation state and binding to 14-3-3.

**The Acidic leucine-rich nuclear phosphoprotein 32 family member A (ANP32A)** gene encodes a multifunctional protein involved in tumor suppression, apoptosis, cell cycle progression or transcription, modulation of histone acetylation and transcription, regulation of mRNA nuclear-to-cytoplasmic translocation and stability by its association with ELAVL1 (Hu-antigen R) [3], regulation of mRNA nuclear-to-cytoplasmic translocation and stability by its association with ELAVL1 (Hu-antigen R) [4]. It plays an essential role in influenza A, B and C viral genome replication [5] and in foamy virus mRNA export from the nucleus [6].

**The C1orf6/Ubiquilin 4 (UBQLN4)** gene encodes a regulator of protein degradation by the proteasome [7]. It is a regulator of DNA repair via MRE11 degradation by the proteasome [8]. UBQLN4 is associated with amyotrophic lateral sclerosis and frontotemporal dementia.

**The Chromosome 1 Open Reading Frame 43 (C1orf43)** encodes a protein coding gene. Gene Ontology annotations related to this gene include oxidoreductase activity, acting on the CH-OH group of donors, NAD or NADP as acceptor and obsolete coenzyme binding. According to Human Protein Atlas a high expression correlates with good prognosis is ccRCC.

**The Chromosome 9 open reading frame 85 (C9orf85)** gene encodes a protein with an as-yet unconfirmed function. However, its expression is notably high in highly differentiated cells [9].

**The cytosolic iron-sulfur protein assembly protein (CIAO1)** gene encodes a protein that forms a complex with FAM96B, MMS19, the co-chaperone HSC20 and the scaffold protein ISCU to assist iron-sulfur cluster incorporation into cytoplasmic and nuclear iron-sulfur proteins [10]. CIAO1 is associated with diseases like Neurodegeneration With Brain Iron Accumulation and Autism Spectrum Disorder.

**The Casein Kinase 1 Alpha 1 (CNSK1A1)** gene encodes a protein kinase that positively regulates mTORC1 and mTORC2 signaling in response to nutrients [11] and inhibits the NLRP3 inflammasome. Given its involvement in diverse cellular, physiological, and pathological processes, CSNK1A1 is considered a promising therapeutic target [12].

**The Translation initiation factor eIF-2B subunit alpha (EIF2B1)** gene encodes a protein that is part of the eIF2B complex, which plays a critical role in translation initiation [13]. Mutations in this gene are associated with leukoencephalopathy.

**The F-Box Protein 7 (FBXO7)** gene encodes a protein that functions as a substrate recognition component of an E3 ubiquitin-protein ligase complex, which mediates the ubiquitination and subsequent proteasomal degradation of target proteins. FBXO7 is involved in regulating processes such as cell cycle progression, cell proliferation, and chromosome stability [14] and has been implicated in Parkinson's disease.

**The Lysine Acetyltransferase 6B (KAT6B)** gene encodes a protein involved in the regulation of transcription either negatively or positively. It may be involved in cortex development and has been associated to Genitopatellar Syndrome and Ohdo Syndrome, Sbbys Variant. Low expression is linked to the prognosis of hepatocellular carcinoma [15].

**The Integrator complex subunit 8 (INTS8)** gene encodes a protein that is part of a complex involved in the transcription of small nuclear RNAs (snRNA) U1 and U2, as well as their 3'-box-dependent processing. Mutations in this gene have been associated with Neurodevelopmental Disorder with Cerebellar Hypoplasia and Spasticity, and Long QT Syndrome. It appears as a therapeutic target for cholangiocarcinoma [16].

**The S-methyl-5'-thioadenosine phosphorylase (MTAP)** gene encodes a phosphorylase that plays a crucial role in polyamine metabolism. Downregulation or deletion of the MTAP gene leads to an accumulation of methyl-5'-thioadenosine (MTA), which promotes tumor growth and significantly enhances the proliferation of tumor cells [17].

**The Mitochondrial translational release factor 1-like (MTRF1L)** gene encodes a nuclear protein that recognizes termination codons and releases mitochondrial ribosomes from the synthesized protein [18]. According to the Human Protein Atlas high expression correlates to a shorter survival in breast liver and lung cancer but to a longer survival in kidney cancers.

**The Nuclear prelamin A recognition factor (NARF)** gene encodes a protein located in the nucleus, where it partially colocalizes with the nuclear lamina. NARF has been associated with Amyotrophic Lateral Sclerosis Type 12 and Acute Retrobulbar Neuritis. NARF is a hypoxia-induced coactivator for OCT4-mediated breast cancer stem cell specification [19].

**The Nucleobindin-1 (NUCB1)** gene encodes a member of a small calcium-binding EF-hand protein family. It is thought to have a key role in Golgi calcium homeostasis and $Ca^{2+}$-regulated signal transduction events. It has been associated with Staphyloenterotoxemia and Cervical Adenomyoma. According to the Human Protein Atlas high expression correlates to a longer survival in pancreatic and kidney cancers [20].

**The Optic atrophy 3 protein (OPA3)** gene encodes a protein that may play some role in mitochondrial processes. Mutations in this gene result in 3-methylglutaconic aciduria type III and autosomal dominant optic atrophy and cataract. Low OPA3 expression is associated with squamous cell carcinoma in paranasal sinus tumors [21]. According to the Human Protein Atlas high expression correlates to a shorter survival in liver cancers but to a longer survival in kidney cancers.

**The Protocadherin gamma-C3 (PCDHGC3)** gene encodes a member This gene is a member of the protocadherin gamma gene cluster. PCDHGC3 is strongly expressed in glioblastoma and its high expression is associated with longer progression-free survival [22].

**The 26S proteasome non-ATPase regulatory subunit 3 (PSMD3)** gene encodes a member of the proteasome subunit S3 family, serving as a non-ATPase component of the 19S regulatory lid within the 26S proteasome complex. PSMD3 plays a role in stabilizing HER2, protecting it from degradation in breast cancer, and is thus associated with poor prognosis in these cancers [23] as well as in various other cancer types [24].

**The Ras-related protein Rab-9A (RAB9A)** gene encodes a small GTPase involved in protein transport between endosomes and the trans Golgi network. RAB9A  has an oncogenic role in liver cancers [25]. According to the Human Protein Atlas high RAB9A expression is associated with shorter survival in glioma and pancreatic cancers but longer survival in colorectal and kidney cancers.

**The Squamous cell carcinoma antigen recognized by T-cells 3 (SART3)** gene encodes an RNA-binding nuclear protein that serves as a tumor-rejection antigen recognized by T-cells [26]. This protein is involved in tumor antigenicity, regulation of gene transcription, mRNA synthesis, stem cell proliferation and differentiation, and embryogenesis [27]. Notably, SART3 regulates IL-8 expression and has been shown to predict clinical outcomes in melanoma [28]. According to the Human Protein Atlas high SART3 expression is associated with shorter survival in colorectal and liver cancers but longer survival in glioma.

**The Superkiller viralicidic activity 2-like 2 (SKIV2L2)/Mtr4 Exosome RNA Helicase (MTREX)** gene encodes an enzyme that catalyzes the ATP-dependent unwinding of RNA duplexes with a single-stranded 3' RNA extension. This helicase is a core subunit in several protein complexes, including TRAMP-like, nuclear exosome targeting (NEXT), and poly(A) tail exosome targeting (PAXT) complexes[29]. MTREX drives liver tumorigenesis by promoting cancer metabolic switch through alternative splicing [30]. According to the Human Protein Atlas, high MTREX expression is linked to shorter survival in ovarian cancer but longer survival in kidney cancer.

**The Interleukin 1 Receptor Associated Kinase 1 Binding Protein 1 (IRAK1BP1*)*/SIMPL** gene encodes a key component of the IRAK1-dependent TNFRSF1A signaling pathway, which activates NF-kappa-B and supports cell survival. IRAK1BP1 has been identified as a prognostic factor in lung adenocarcinoma [31]. According to the Human Protein Atlas, high expression of IRAK1BP1 is associated with longer survival in colorectal cancer.

**The SMU1 DNA Replication Regulator And Spliceosomal Factor (SMU1)** gene encodes a spliceosomal component involved in regulating alternative splicing, essential for proper mitotic spindle assembly and normal progression through mitosis [32]. SMU1 Knockdown suppresses gastric carcinoma growth [33]. According to the Human Protein Atlas, high SMU1 expression is linked to shorter survival in liver cancer but longer survival in kidney cancer.

**The TBC1 domain family member 1 (TBC1D1)** gene encodes the founding member of a protein family characterized by a 180- to 200-amino acid TBC domain, believed to play a role in regulating cell growth and differentiation. Members of the TBC family function as GTPase-activating proteins (GAPs) for small GTPases, contributing to cell cycle regulation. TBC1D1 has been identified as a novel prognostic biomarker and is associated with resistance to immunotherapy in gliomas [34]. According to the Human Protein Atlas, high TBC1D1 expression is linked to longer survival in pancreatic and kidney cancer.

**The Transmembrane Protein 167B (TMEM167B)** gene encodes a protein located in the Golgi apparatus, although its function remains unknown. According to the Human Protein Atlas, high TMEM167B expression is associated with shorter survival in liver and urothelial cancers but with longer survival in kidney cancer.

**The Ubiquitin-associated protein 2-like (UBAP2L)** gene encodes a protein that recruits the ubiquitination machinery to RNA polymerase II for polyubiquitination, removal and degradation, when the nucleotide excision repair machinery fails to resolve DNA damage [35]. UBAP2L has a crucial involvement in various cellular processes such as cell cycle regulation, stem cell activity and stress-response signaling. In addition, UBAP2L has recently emerged as a master regulator of growth and proliferation in several human cancers, where it is suggested to display oncogenic properties [36]. UBAP2L enhances growth and metastasis of gastric cancer cells [37]. According to the Human Protein Atlas, high UBAP2L expression is associated with shorter survival in liver and kidney cancer.

**The Ubiquitin-conjugating enzyme E2 G2 (UBE2G2)** gene encodes and enzymes that catalyzes covalent attachment of ubiquitin to other proteins [38]. According to the Human Protein Atlas, high UBE2G2 expression is associated with longer survival in kidney cancer.

**The Ubiquitin Conjugating Enzyme E2 Q1 (UBE2Q1)** gene encodes an enzyme that catalyzes the covalent attachment of ubiquitin to target proteins. UBE2Q1 has been identified as a downregulated gene in pediatric acute lymphoblastic leukemia [39] and is considered a potential prognostic marker for high-grade serous ovarian cancer [40].

**The Ubiquitin-conjugating enzyme E2 R2 (UBE2R2)** gene encodes an enzyme that catalyzes the covalent attachment of ubiquitin to target proteins. UBE2R2 is phosphorylated by casein kinase 2 to regulates beta catenin degradation [41]. According to the Human Protein Atlas, high UBE2R2 expression is associated with shorter survival in colorectal, liver, lung and pancreatic cancers but with longer survival in kidney cancer.

**The Zinc Finger Protein 407 (ZNF407/ FLJ13839)** encodes a zinc finger protein with an unknown function. Mutations in this gene have been associated with several conditions, including short stature, impaired intellectual development, microcephaly, hypotonia, ocular anomalies, and Developmental Malformations-Deafness-Dystonia Syndrome.

**The Zinc Finger Protein 585A (ZNF585A)** gene encodes a predicted DNA-binding protein that regulates RNA polymerase II, though its specific functions have not yet been described.

**References**


[1] Yang H, Jiang X, Li B, Yang HJ, Miller M, Yang A, et al. Mechanisms of mTORC1 activation by RHEB and inhibition by PRAS40. Nature. 2017;552:368-73.

[2] Wang L, Harris TE, Lawrence JC, Jr. Regulation of proline-rich Akt substrate of 40 kDa (PRAS40) function by mammalian target of rapamycin complex 1 (mTORC1)-mediated phosphorylation. J Biol Chem. 2008;283:15619-27.

[3] Santa-Coloma TA. Anp32e (Cpd1) and related protein phosphatase 2 inhibitors. Cerebellum. 2003;2:310-20.

[4] Mazroui R, Di Marco S, Clair E, von Roretz C, Tenenbaum SA, Keene JD, et al. Caspase-mediated cleavage of HuR in the cytoplasm contributes to pp32/PHAP-I regulation of apoptosis. J Cell Biol. 2008;180:113-27.

[5] Wei X, Liu Z, Wang J, Yang R, Yang J, Guo Y, et al. The interaction of cellular protein ANP32A with influenza A virus polymerase component PB2 promotes vRNA synthesis. Arch Virol. 2019;164:787-98.

[6] Bodem J. Regulation of foamy viral transcription and RNA export. Adv Virus Res. 2011;81:1-31.

[7] Suzuki R, Kawahara H. UBQLN4 recognizes mislocalized transmembrane domain proteins and targets these to proteasomal degradation. EMBO Rep. 2016;17:842-57.

[8] Jachimowicz RD, Beleggia F, Isensee J, Velpula BB, Goergens J, Bustos MA, et al. UBQLN4 Represses Homologous Recombination and Is Overexpressed in Aggressive Tumors. Cell. 2019;176:505-19 e22.

[9] Chen BZ, Yu SL, Singh S, Kao LP, Tsai ZY, Yang PC, et al. Identification of microRNAs expressed highly in pancreatic islet-like cell clusters differentiated from human embryonic stem cells. Cell Biol Int. 2011;35:29-37.

[10] Kim KS, Maio N, Singh A, Rouault TA. Cytosolic HSC20 integrates de novo iron-sulfur cluster biogenesis with the CIAO1-mediated transfer to recipients. Hum Mol Genet. 2018;27:837-52.

[11] Gao D, Inuzuka H, Tan MK, Fukushima H, Locasale JW, Liu P, et al. mTOR drives its own activation via SCF(betaTrCP)-dependent degradation of the mTOR inhibitor DEPTOR. Mol Cell. 2011;44:290-303.

[12] Jiang S, Zhang M, Sun J, Yang X. Casein kinase 1alpha: biological mechanisms and theranostic potential. Cell Commun Signal. 2018;16:23.

[13] Sekine Y, Zyryanova A, Crespillo-Casado A, Fischer PM, Harding HP, Ron D. Stress responses. Mutations in a translation initiation factor identify the target of a memory-enhancing compound. Science. 2015;348:1027-30.

[14] Hsu JM, Lee YC, Yu CT, Huang CY. Fbx7 functions in the SCF complex regulating Cdk1-cyclin B-phosphorylated hepatoma up-regulated protein (HURP) proteolysis by a proline-rich region. J Biol Chem. 2004;279:32592-602.

[15] Jiang J, Wang HJ, Mou XZ, Zhang H, Chen Y, Hu ZM. Low Expression of KAT6B May Affect Prognosis in Hepatocellular Carcinoma. Technol Cancer Res Treat. 2021;20:15330338211033063.

[16] Zhou Q, Ji L, Shi X, Deng D, Guo F, Wang Z, et al. INTS8 is a therapeutic target for intrahepatic cholangiocarcinoma via the integration of bioinformatics analysis and experimental validation. Sci Rep. 2021;11:23649.

[17] Kirovski G, Stevens AP, Czech B, Dettmer K, Weiss TS, Wild P, et al. Down-regulation of methylthioadenosine phosphorylase (MTAP) induces progression of hepatocellular

carcinoma via accumulation of 5'-deoxy-5'-methylthioadenosine (MTA). Am J Pathol. 2011;178:1145-52.

[18] Nozaki Y, Matsunaga N, Ishizawa T, Ueda T, Takeuchi N. HMRF1L is a human mitochondrial translation release factor involved in the decoding of the termination codons UAA and UAG. Genes Cells. 2008;13:429-38.

[19] Yang Y, Chen C, Zuo Q, Lu H, Salman S, Lyu Y, et al. NARF is a hypoxia-induced coactivator for OCT4-mediated breast cancer stem cell specification. Sci Adv. 2022;8:eabo5000.

[20] Hua YQ, Zhang K, Sheng J, Ning ZY, Li Y, Shi WD, et al. NUCB1 Suppresses Growth and Shows Additive Effects With Gemcitabine in Pancreatic Ductal Adenocarcinoma via the Unfolded Protein Response. Front Cell Dev Biol. 2021;9:641836.

[21] Yang Z, Zhang Y, Wang X, Huang J, Guo W, Wei P, et al. Putative biomarkers of malignant transformation of sinonasal inverted papilloma into squamous cell carcinoma. J Int Med Res. 2019;47:2371-80.

[22] Feldheim J, Wend D, Lauer MJ, Monoranu CM, Glas M, Kleinschnitz C, et al. Protocadherin Gamma C3 (PCDHGC3) Is Strongly Expressed in Glioblastoma and Its High Expression Is Associated with Longer Progression-Free Survival of Patients. Int J Mol Sci. 2022;23.

[23] Fararjeh AS, Chen LC, Ho YS, Cheng TC, Liu YR, Chang HL, et al. Proteasome 26S Subunit, non-ATPase 3 (PSMD3) Regulates Breast Cancer by Stabilizing HER2 from Degradation. Cancers (Basel). 2019;11.

[24] Rubio AJ, Bencomo-Alvarez AE, Young JE, Velazquez VV, Lara JJ, Gonzalez MA, et al. 26S Proteasome Non-ATPase Regulatory Subunits 1 (PSMD1) and 3 (PSMD3) as Putative Targets for Cancer Prognosis and Therapy. Cells. 2021;10.

[25] Sun P, Li L, Li Z. RAB9A Plays an Oncogenic Role in Human Liver Cancer Cells. Biomed Res Int. 2020;2020:5691671.

[26] Harada K, Yamada A, Yang D, Itoh K, Shichijo S. Binding of a SART3 tumor-rejection antigen to a pre-mRNA splicing factor RNPS1: a possible regulation of splicing by a complex formation. Int J Cancer. 2001;93:623-8.

[27] Whitmill A, Timani KA, Liu Y, He JJ. Tip110: Physical properties, primary structure, and biological functions. Life Sci. 2016;149:79-95.

[28] Timani KA, Gyorffy B, Liu Y, Mohammad KS, He JJ. Tip110/SART3 regulates IL-8 expression and predicts the clinical outcomes in melanoma. Mol Cancer. 2018;17:124.

[29] Lubas M, Christensen MS, Kristiansen MS, Domanski M, Falkenby LG, Lykke-Andersen S, et al. Interaction profiling identifies the human nuclear exosome targeting complex. Mol Cell. 2011;43:624-37.

[30] Yu L, Kim J, Jiang L, Feng B, Ying Y, Ji KY, et al. MTR4 drives liver tumorigenesis by promoting cancer metabolic switch through alternative splicing. Nat Commun. 2020;11:708.

[31] Guo L, Zhou W, Xu Z, Cao X, Wan S, Zhang YY, et al. Identification of IRAK1BP1 as a candidate prognostic factor in lung adenocarcinoma. Front Oncol. 2023;13:1132811.

[32] Bertram K, Agafonov DE, Dybkov O, Haselbach D, Leelaram MN, Will CL, et al. Cryo-EM Structure of a Pre-catalytic Human Spliceosome Primed for Activation. Cell. 2017;170:701-13 e11.

[33] Qian M, Liang X, Zeng Q, Zhang C, He N, Ma J. SMU1 Knockdown Suppresses Gastric Carcinoma Growth, Migration, and Invasion and Modulates the Cell Cycle. Cancer Control. 2024;31:10732748241281716.

[34] Song D, Yang Q, Li L, Wei Y, Zhang C, Du H, et al. Novel prognostic biomarker TBC1D1 is associated with immunotherapy resistance in gliomas. Front Immunol. 2024;15:1372113.

[35] Herlihy AE, Boeing S, Weems JC, Walker J, Dirac-Svejstrup AB, Lehner MH, et al. UBAP2/UBAP2L regulate UV-induced ubiquitylation of RNA polymerase II and are the human orthologues of yeast Def1. DNA Repair (Amst). 2022;115:103343.

[36] Guerber L, Pangou E, Sumara I. Ubiquitin Binding Protein 2-Like (UBAP2L): is it so NICE After All? Front Cell Dev Biol. 2022;10:931115.

[37] Lin S, Yan Z, Tang Q, Zhang S. Ubiquitin-associated protein 2 like (UBAP2L) enhances growth and metastasis of gastric cancer cells. Bioengineered. 2021;12:10232-45.

[38] David Y, Ziv T, Admon A, Navon A. The E2 ubiquitin-conjugating enzymes direct polyubiquitination to preferred lysines. J Biol Chem. 2010;285:8595-604.

[39] Seghatoleslam A, Bozorg-Ghalati F, Monabati A, Nikseresht M, Owji AA. UBE2Q1, as a Down Regulated Gene in Pediatric Acute Lymphoblastic Leukemia. Int J Mol Cell Med. 2014;3:95-101.

[40] Topno R, Singh I, Kumar M, Agarwal P. Integrated bioinformatic analysis identifies UBE2Q1 as a potential prognostic marker for high grade serous ovarian cancer. BMC Cancer. 2021;21:220.

[41] Semplici F, Meggio F, Pinna LA, Oliviero S. CK2-dependent phosphorylation of the E2 ubiquitin conjugating enzyme UBC3B induces its interaction with beta-TrCP and enhances beta-catenin degradation. Oncogene. 2002;21:3978-87.

#### Legends to supplemental figure and tables

**Supplementary Fig. 1: Genes Associated with Longer OS/DFS by Age Group**

This figure illustrates the expression levels of genes linked to longer OS and DFS, stratified by age tranches. The genes in the prognostic signature are displayed across different age groups. Statistical significance between the age groups is highlighted, with P-values from the Kruskal-Wallis's test provided for each gene and Wilcoxon rank sum test for comparison between age groups.

**Supplementary Fig. 2: Genes Associated with Shorter OS/DFS by Age Group**

This figure illustrates the expression levels of genes linked to shorter OS and DFS, stratified by age tranches. The genes in the prognostic signature are displayed across different age groups. Statistical significance between the age groups is highlighted, with P-values derived from the Kruskal-Wallis' test provided for each gene and Wilcoxon rank sum test for comparison between age groups.

**Supplementary Fig. 3: Data Quality Assessment.**

Principal component analysis (PCA) was performed on the selected genes for OS (A, B) and DFS (C, D). Panels A and C show the percentage of variance explained by each PCA dimension. Panels B and D display the distribution of patients in the Euclidean plane, based on the two most important dimensions (1 and 2), to assess patient clustering and identify potential outliers.

**Supplementary Fig. 4: Variable Importance (24 variables) in the random survival forest model for OS, M0 patients.** Model parameters were sample size = 294, number of deaths = 59, number of trees = 2000, forest terminal node size = 12, number of variables tried at each split = 3 and (OOB) requested performance error = 0.289. Clinical parameters are presented on a black background.

This figure displays the importance of 24 variables used in the random survival forest model for OS prediction. Variables with higher values have a greater impact on the final prognostic score. Notably, the UBE2G2 gene (highlighted in red) stands out as the most statistically significant, second most influential variable, exerting the strongest effect on the model's predictions.

**Supplementary Fig. 5: Variable Importance (23 variables) in the random survival forest model for PFS, M0 patients** Model parameters were sample size = 249, number of events = 34, number of trees = 100, forest terminal node size = 1, number of variables tried at each split = 5 and (OOB) requested performance error = 0.267. This figure displays the importance of 23 variables used in the random survival forest model for DFS prediction. Variables with higher values have a greater impact on the final prognostic score. The SART3 gene stands out as the most influential variable, exerting the strongest effect on the model's predictions.

**Supplementary Fig. 6: Determination of OS of Patients Across Different Age Groups According to Our Model.** Kaplan-Meier analysis of OS in patients stratified by age groups. P-values from Cox regression are presented.

**Supplementary Fig. 7: Determination of DFS of Patients Across Different Age Groups According to Our Model.** Kaplan-Meier analysis of DFS in patients stratified by age groups. P-values from Cox regression are presented. LogRank test was performed in place of Cox bivariate regression when p-value and hazard ration were not calculable.

**Supplementary Fig. 8: Data Quality Assessment.** Principal component analysis (PCA) was performed on the selected genes for OS (A, B) and PFS (C, D). Panels A and C show the percentage of variance explained by each PCA dimension. Panels B and D display the distribution of patients in the Euclidean plane, based on the two most important dimensions (1 and 2), to assess patient clustering and identify potential outliers.

**Supplementary Fig. 9: Variable Importance (24 variables) in the random survival forest model for OS, M1 patients.** Model parameters were sample size = 83, number of deaths = 65, number of trees = 100, forest terminal node size = 1, number of variables tried at each split = 20 and (OOB) requested performance error = 0.384. Clinical parameters are presented on a black background. This figure displays the importance of 24 variables used in the random survival forest model for OS prediction. Variables with higher values have a greater impact on the final prognostic score. Notably, the RAB9A and the PCDHGC3 genes (highlighted in red) stand out as statistically significant, exerting the strongest effect on the model's predictions.

**Supplementary Fig. 10: Variable Importance (23 variables) in the random survival forest model for DFS, M1 patients.** Model parameters were sample size = 56, number of events = 41, number of trees = 100, forest terminal node size = 1, number of variables tried at each split = 20 and (OOB) requested performance error = 0.494. This figure displays the importance of 23 variables used in the random survival forest model for PFS prediction. Variables with higher values have a greater impact on the final prognostic score. The UBA2PL gene stands out as the most influential variable, exerting the strongest effect on the model's predictions.

**Supplementary Fig. 11: Determination of OS of M1 Patients Across Different Age Groups According to Our Model.** Kaplan-Meier analysis of OS in patients stratified by age groups. P-values from Cox regression are presented. LogRank test was performed in place of Cox bivariate regression when p-value and hazard ration were not calculable.

**Supplementary Fig. 12: Determination of DFS of Patients Across Different Age Groups According to Our Model.** Kaplan-Meier analysis of DFS in patients stratified by age groups. P-values from Cox regression proportional are presented.

**Supplementary Table 1: Patient demographics of the TCGA cohort of M0 ccRCC patients.**

**Supplementary Table 2: Correlation Between Clinical Parameters and OS.**

Various clinical parameters—such as tumor stage, tumor size (PT), and Fuhrman grade—were analyzed for their correlation with OS. The table presents the number of patients (n), hazard ratios (HR), 95% confidence intervals (CI), and P-values for each parameter.

**Supplementary Table 3: Correlation Between Clinical Parameters and DFS.**

Various clinical parameters—including tumor stage, tumor size (PT), sex, and Fuhrman grade—were analyzed for their correlation with OS. The table provides the number of patients (headcounts), hazard ratios (HR), 95% confidence intervals (CI 95%), and P-values for each parameter.

**Supplementary Table 4: Parameters of the simplified Cox regression model for OS**: Hazard ratio (low and high) and P values are indicated.

**Supplementary Table 5: Parameters of the simplified Cox regression model for DFS**: Hazard ratio (low and high) and P values are indicated.

**Supplementary Table 6: Patient demographics of the TCGA cohort of M1 ccRCC patients.**

### Figure S1: Pace-Locos T, *et al*.

LONGER OS/DFS

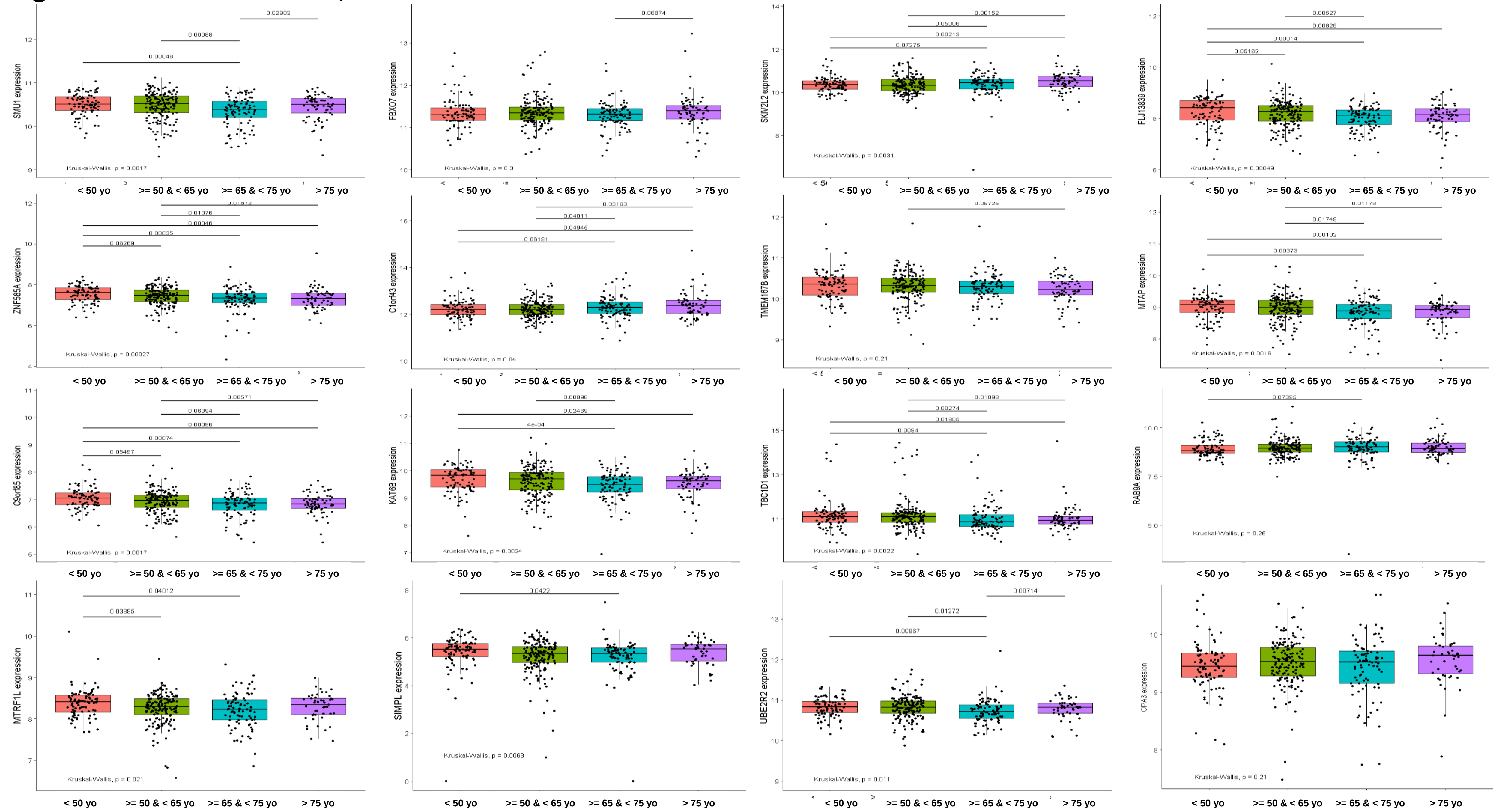

### SHORTER OS/DFS

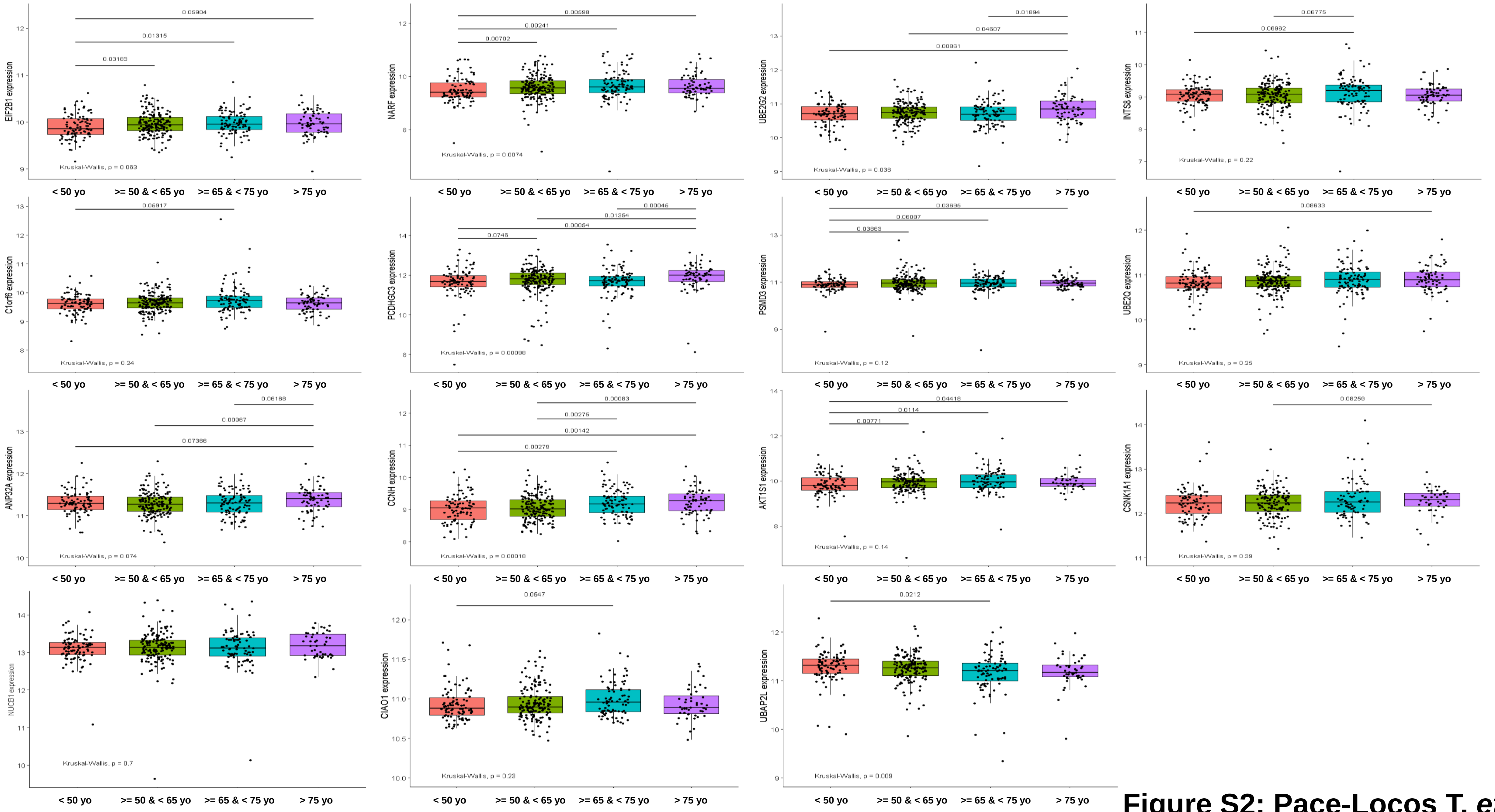


**Figure S2: Pace-Locos T, *et al.***

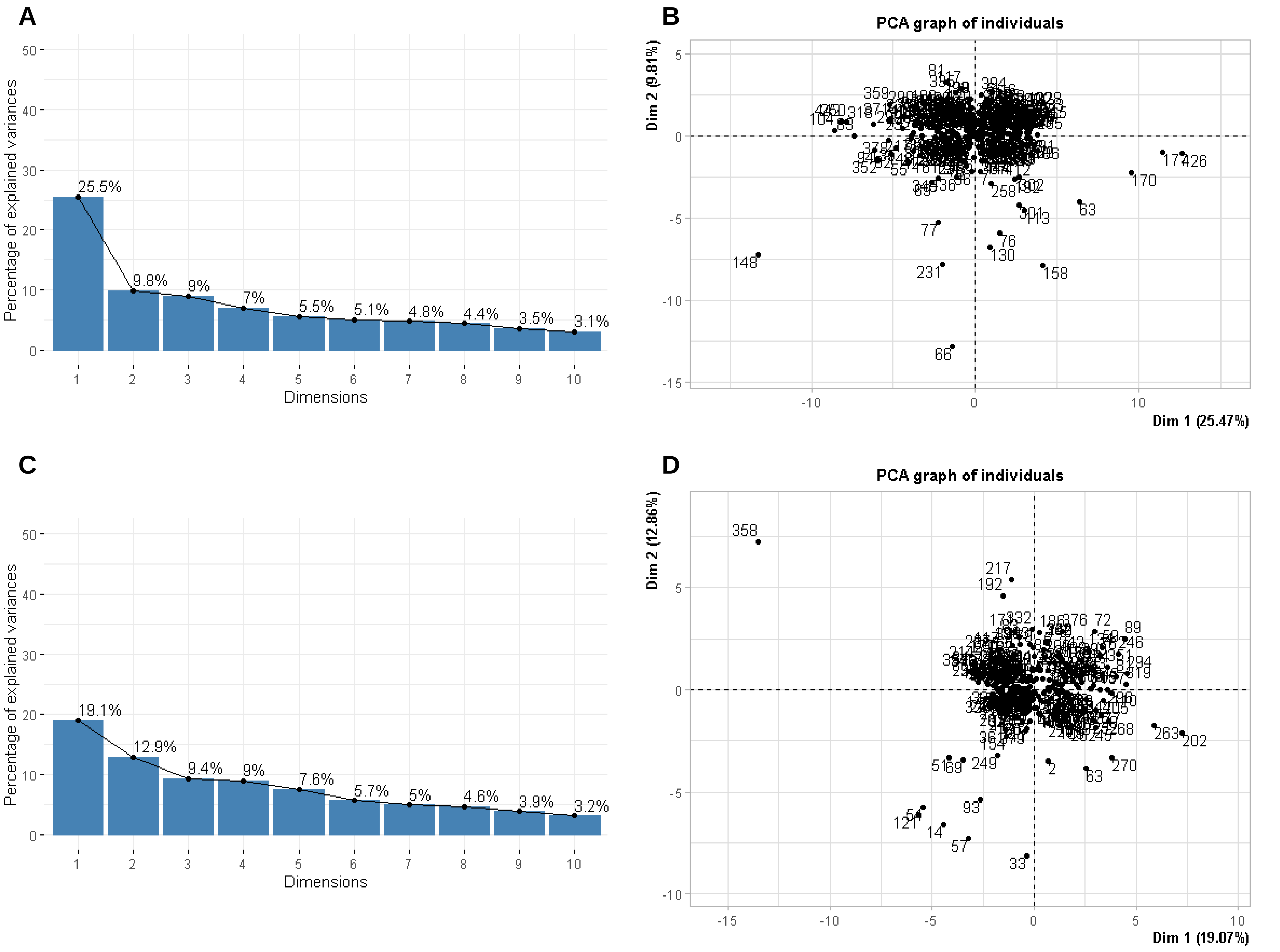


**Supplementary Fig. 3: Pace-Locos T, *et al*.**

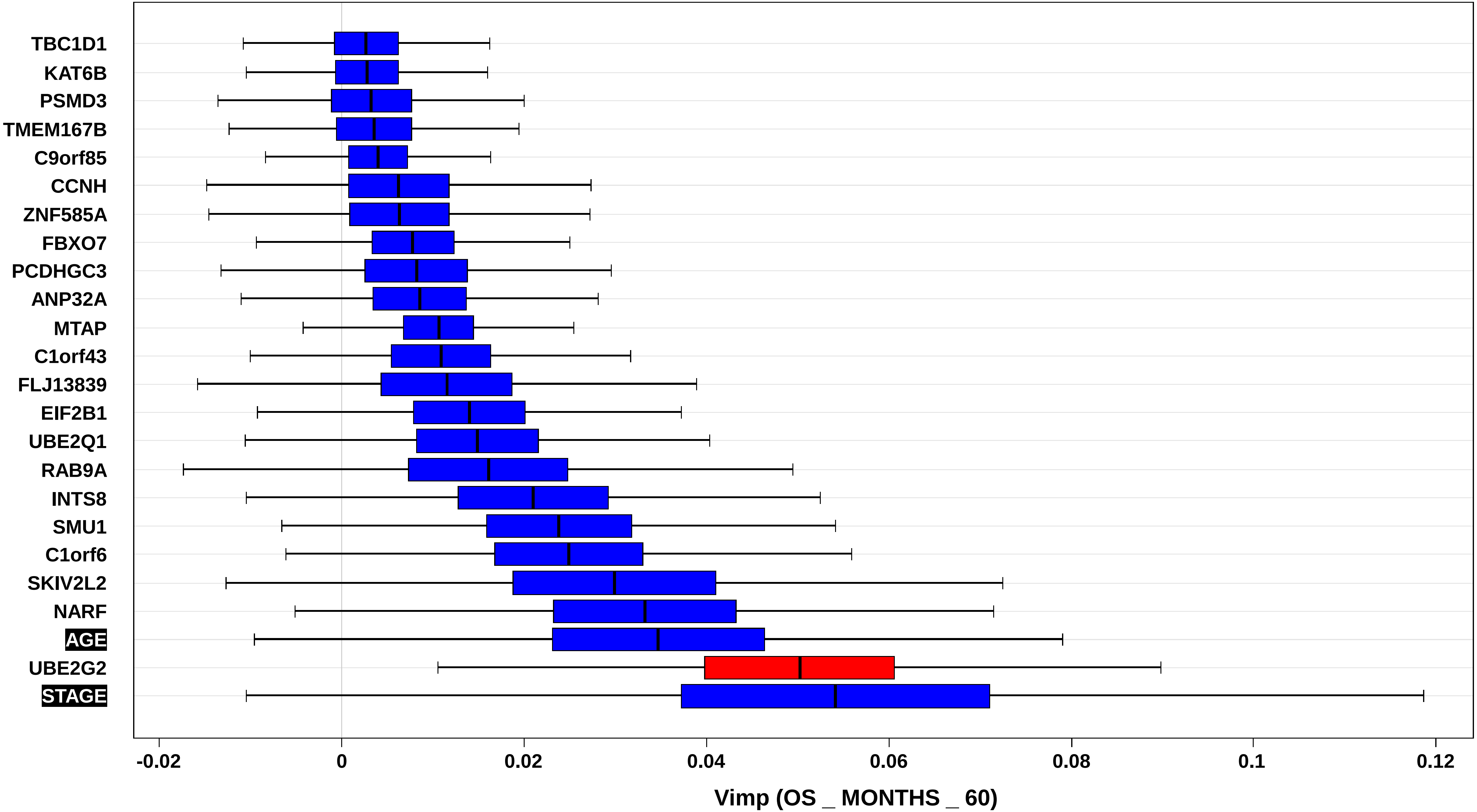


**Supplementary Figure 4: Pace-Locos T, *et al*.**

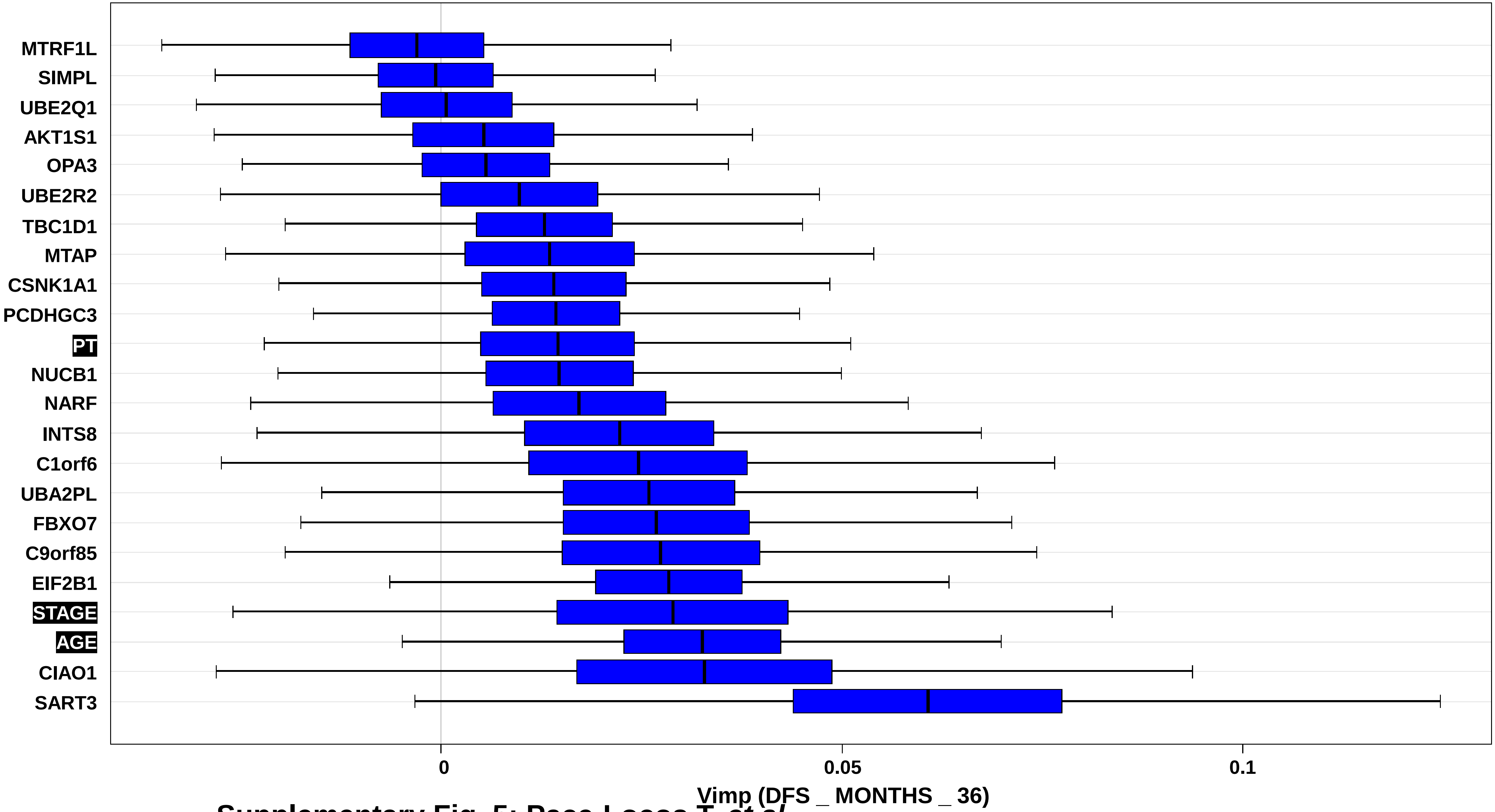


**Supplementary Fig. 5: Pace-Locos T, *et al*.**

#### POP < 50 YO

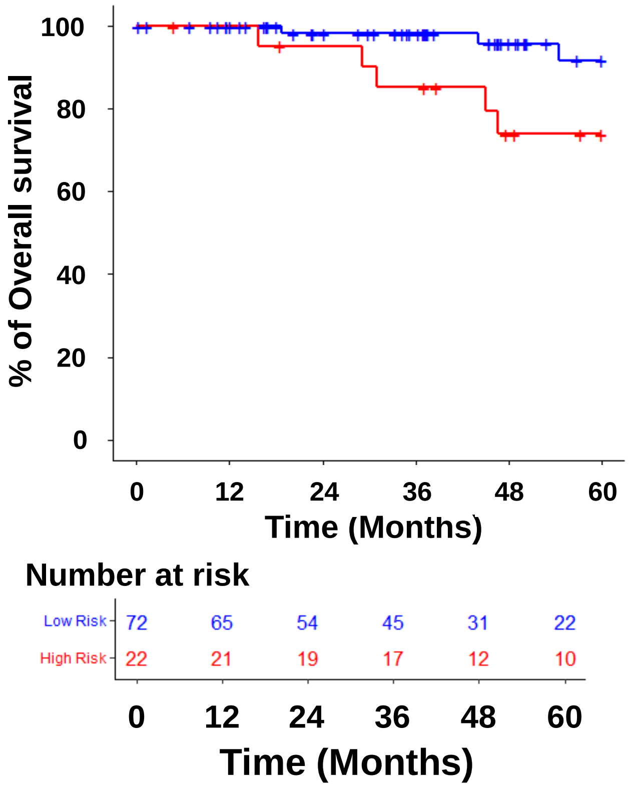


| Variable | Modality | Headcount | HR | CI95% | p-Cox |
|---|---|---|---|---|---|
| Risk groups | | | | | |
| | Low Risk | 72 | 1 | REF | |
| | High Risk | 22 | 4.4 | [1 - 18] | 0.044 |

#### POP >= 50 & < 65 YO

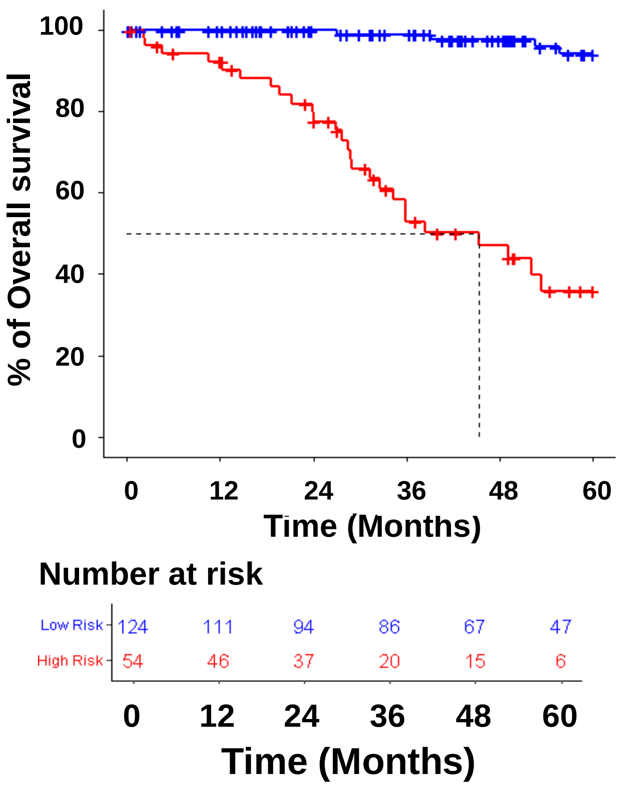


| Variable | Modality | Headcount | HR | CI95% | p-Cox |
|---|---|---|---|---|---|
| Risk groups | | | | | |
| | Low Risk | 124 | 1 | REF | |
| | High Risk | 54 | 22 | [7.6 - 63] | < 0.0001 |

#### POP >= 65 & < 75 YO

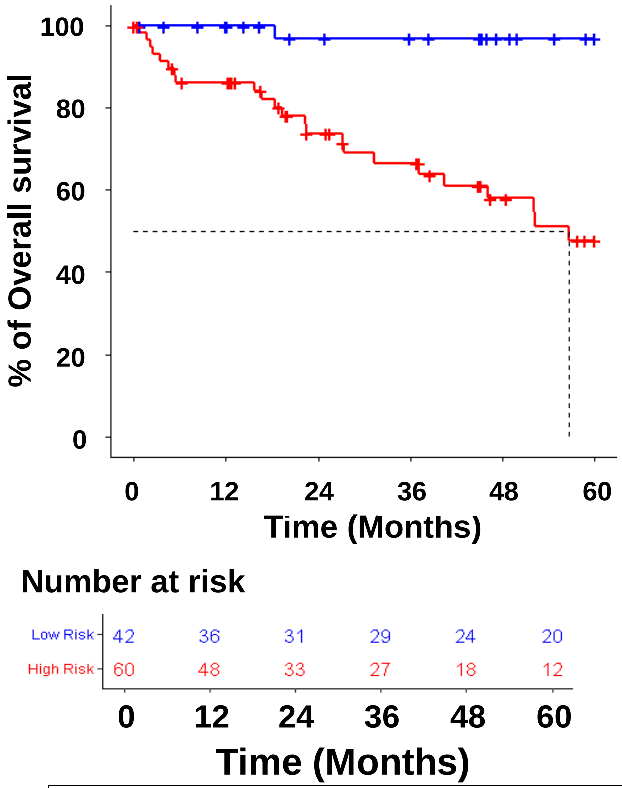


| Variable | Modality | Headcount | HR | CI95% | p-Cox |
|---|---|---|---|---|---|
| Risk groups | | | | | |
| | Low Risk | 42 | 1 | REF | |
| | High Risk | 60 | 21 | [2.9 - 160] | 0.0028 |

#### POP >= 75 YO

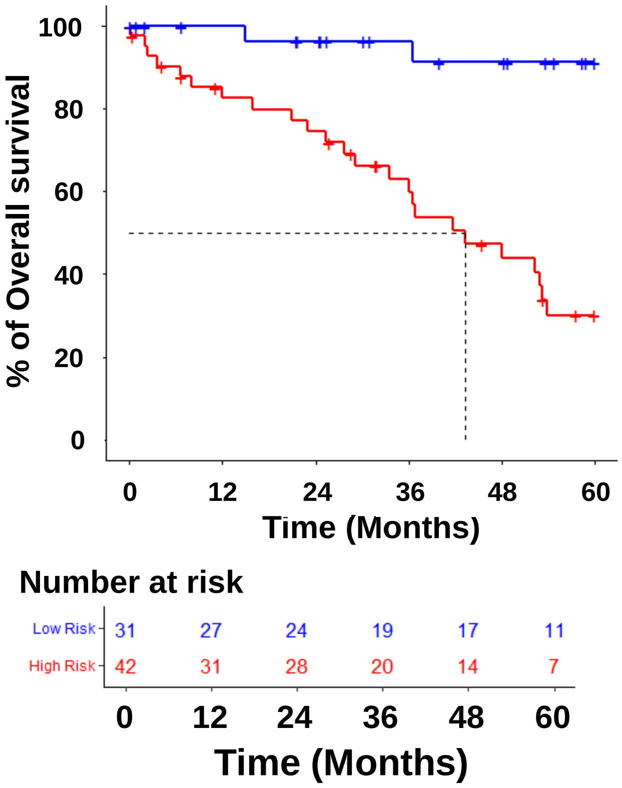


| Variable | Modality | Headcount | HR | CI95% | p-Cox |
|---|---|---|---|---|---|
| Risk groups | | | | | |
| | Low Risk | 31 | 1 | REF | |
| | High Risk | 42 | 11 | [2.7- 48] | < 0.0001 |

**Supplementary Fig. 6: Pace-Locos T, *et al*.**

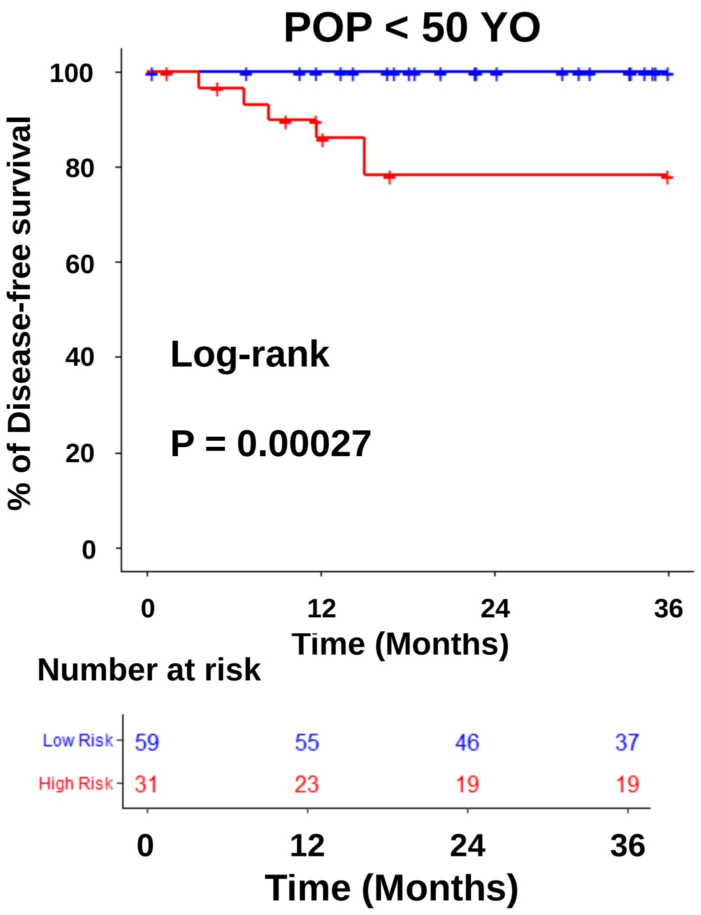


| Variable | Modality | Headcount | HR | CI95% |
|---|---|---|---|---|
| Risk groups | | | | |
| | Low Risk | 31 | 1 | REF |
| | High Risk | 42 | 1.9e+09 | [0- Inf] |

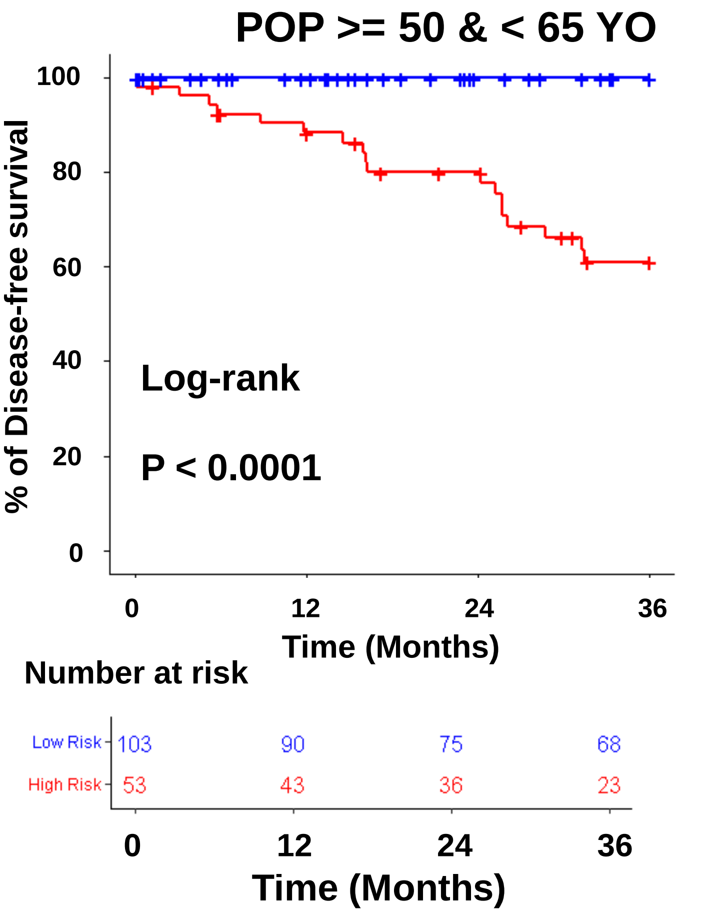


| Variable | Modality | Headcount | HR | CI95% | p-Cox |
|---|---|---|---|---|---|
| Risk groups | | | | | |
| | Low Risk | 103 | 1 | REF | |
| | High Risk | 53 | 1.8e+9 | [0 - Inf] | 1 |

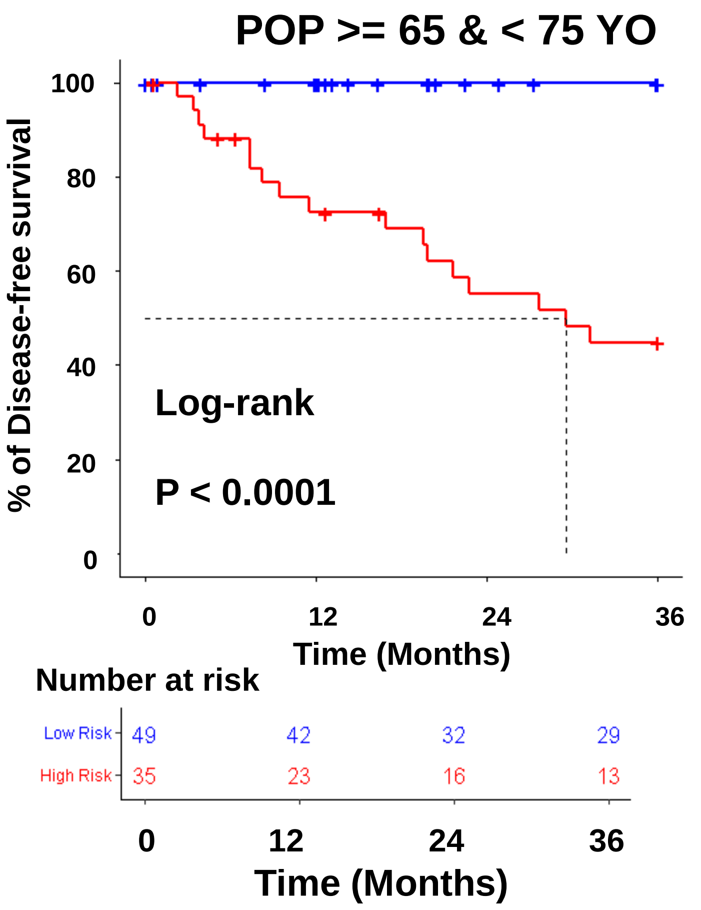


| Variable | Modality | Headcount | HR | CI95% | p-Cox |
|---|---|---|---|---|---|
| Risk groups | | | | | |
| | Low Risk | 103 | 1 | REF | |
| | High Risk | 53 | 1.2e+9 | [0 - Inf] | 1 |

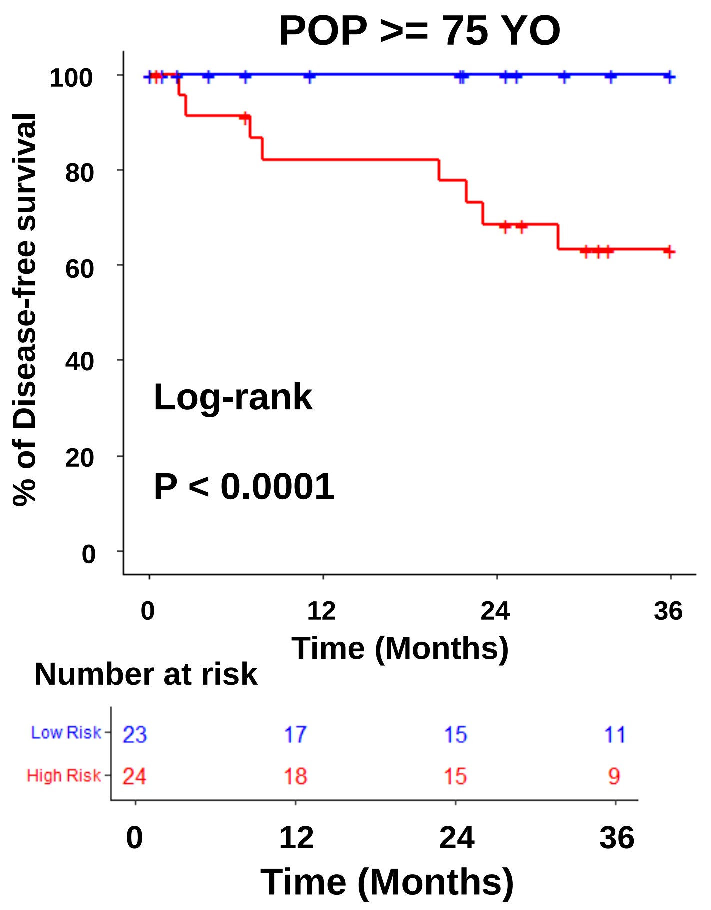


| Variable | Modality | Headcount | HR | CI95% |
|---|---|---|---|---|
| Risk groups | | | | |
| | Low Risk | 23 | 1 | REF |
| | High Risk | 24 | 5.5e+08 | [0- Inf] |

**Supplementary Fig. 7: Pace-Locos T, *et al*.**

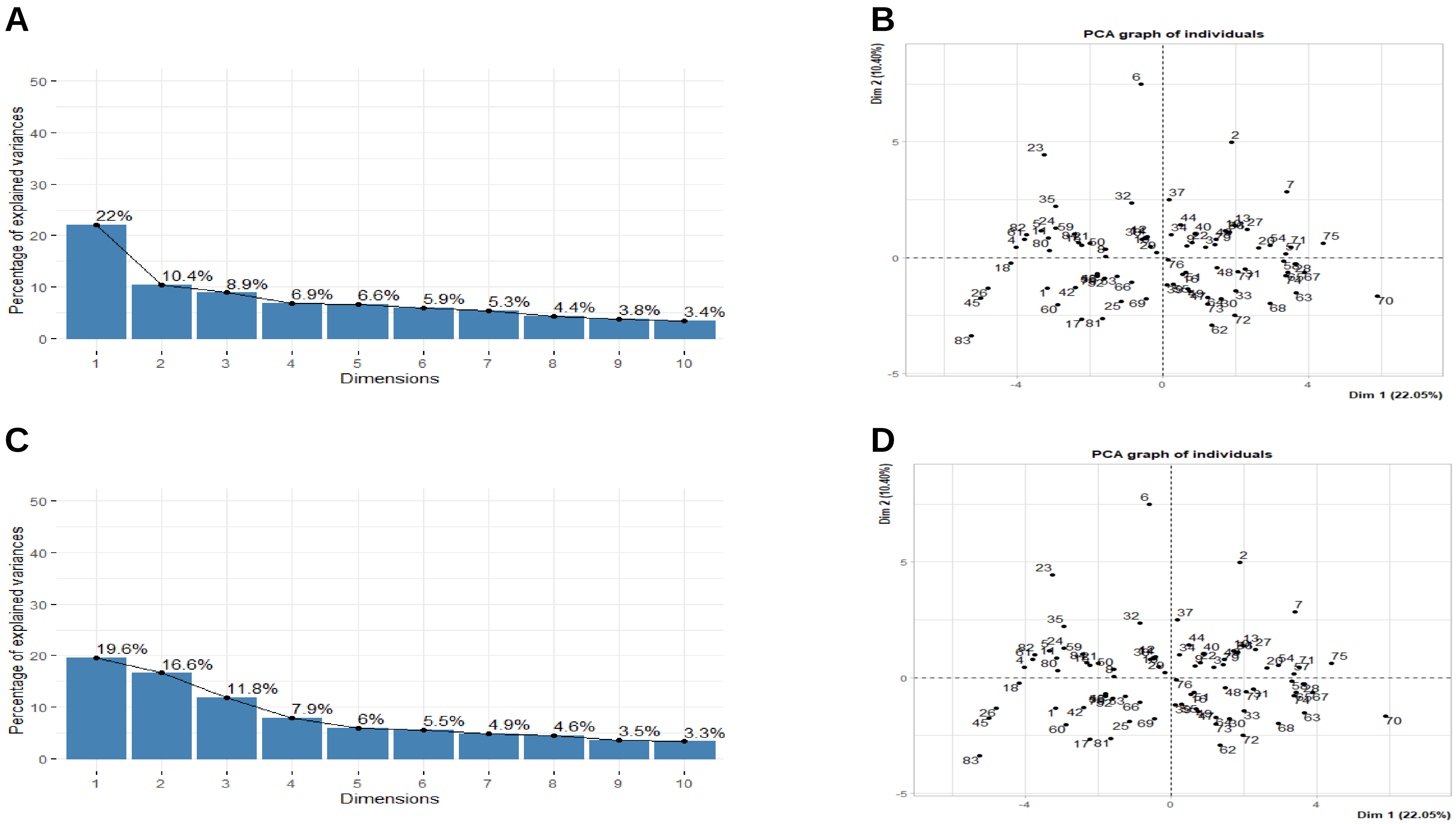


**Supplementary Fig. 8: Pace-Locos T, *et al*.**

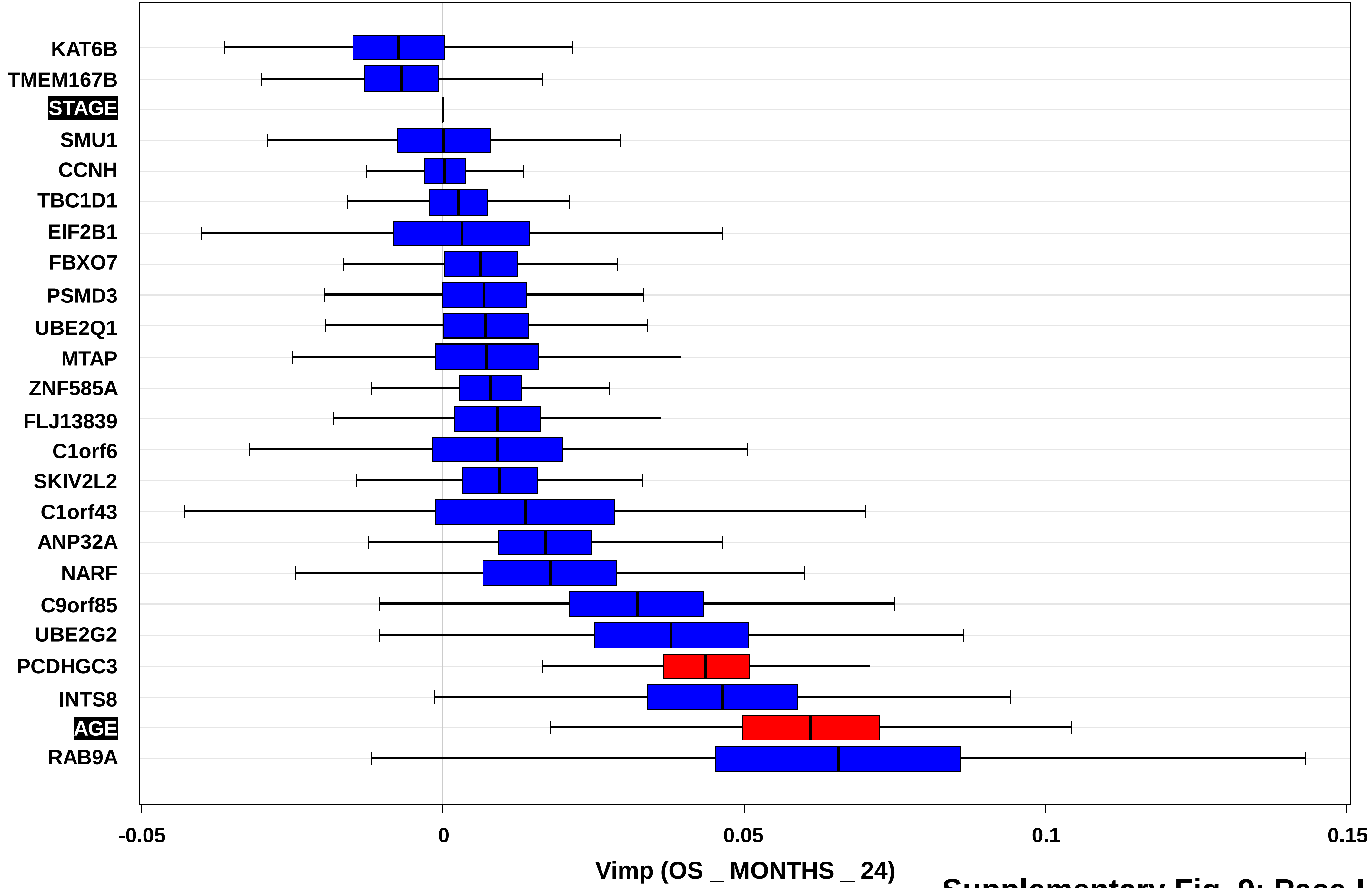


**Supplementary Fig. 9: Pace-Locos T, *et al*.**

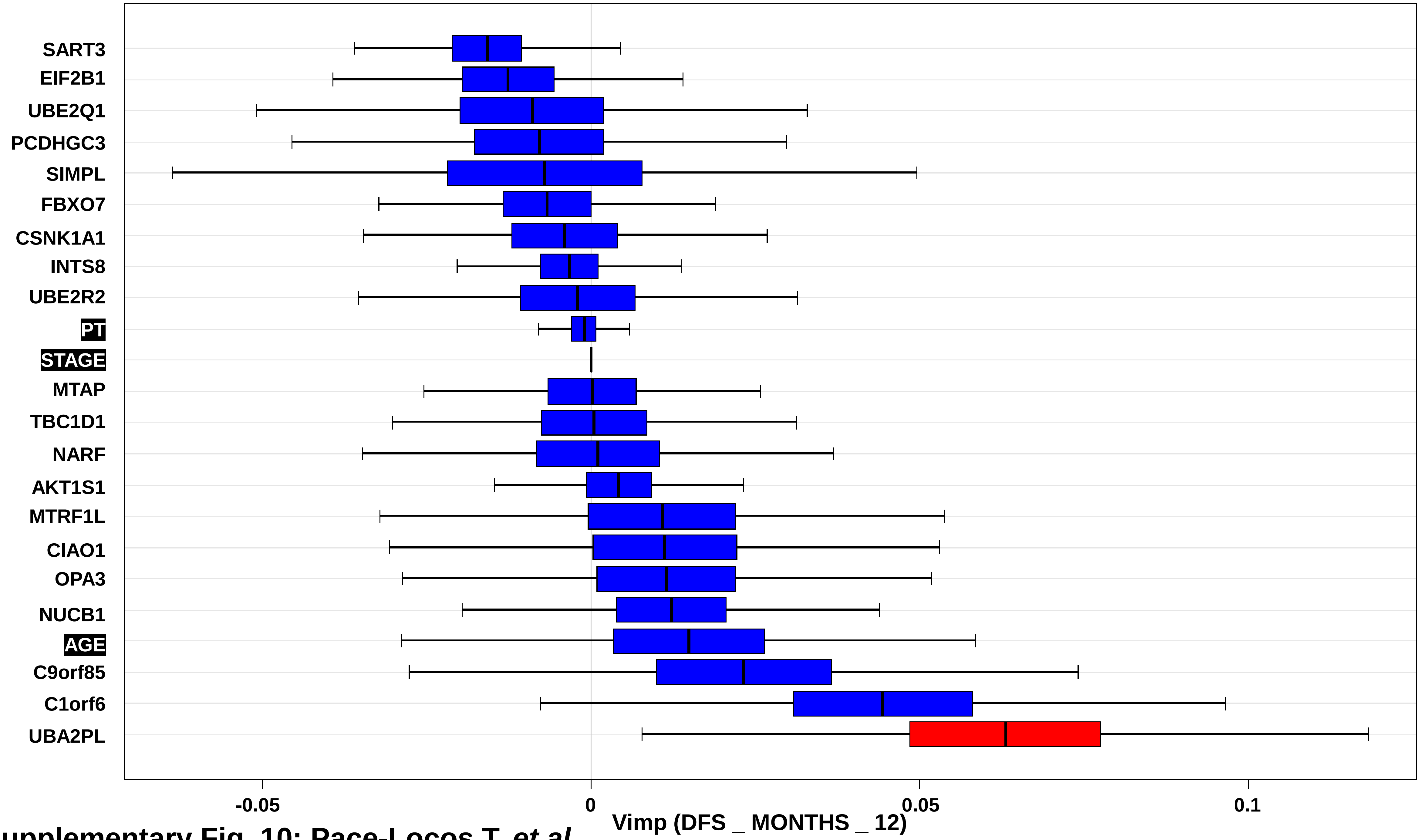


**Supplementary Fig. 10: Pace-Locos T, *et al*.**

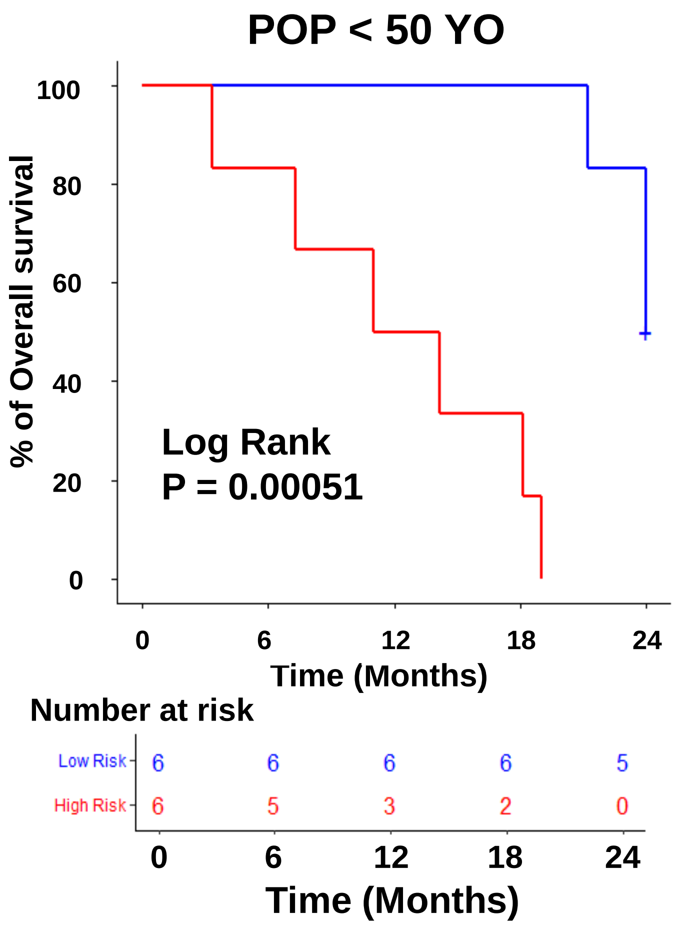


| Variable | Modality | Headcount | HR | CI95% | p-Cox |
|---|---|---|---|---|---|
| Risk groups | | | | | |
| | Low Risk | 6 | 1 | REF | |
| | High Risk | 6 | 4.9e+9 | [0-Inf] | 1 |

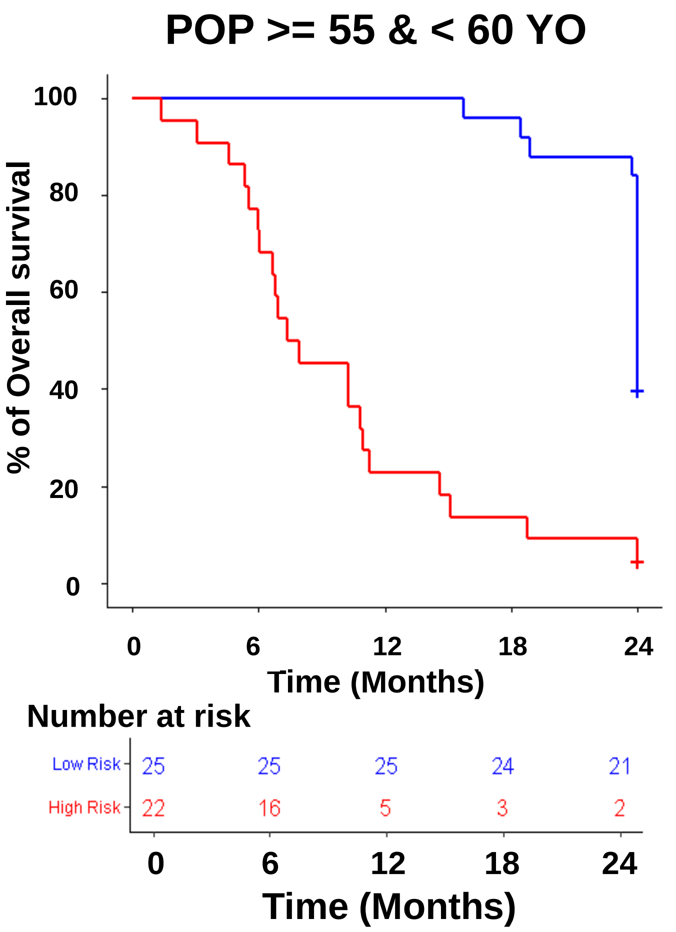


| Variable | Modality | Headcount | HR | CI95% | p-Cox |
|---|---|---|---|---|---|
| Risk groups | | | | | |
| | Low Risk | 25 | 1 | REF | |
| | High Risk | 22 | 6.7 | [3.3- 14] | < 0.0001 |

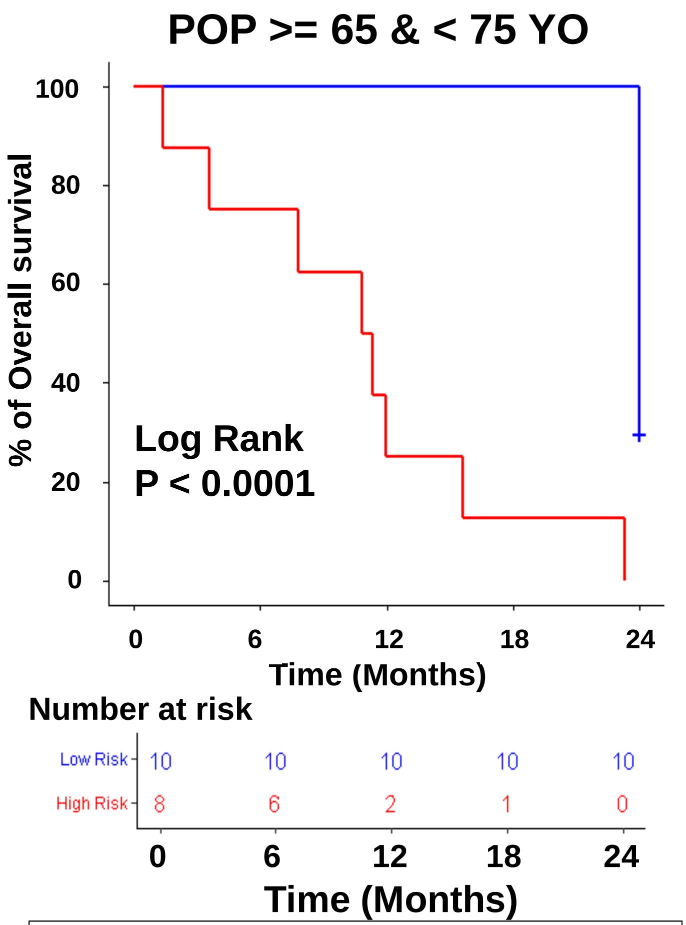


| Variable | Modality | Headcount | HR | CI95% | p-Cox |
|---|---|---|---|---|---|
| Risk groups | | | | | |
| | Low Risk | 23 | 1 | REF | |
| | High Risk | 19 | 45 | [9.8- 210] | < 0.0001 |

**Supplementary Fig. 11: Pace-Locos T, *et al*.**

#### POP < 50 YO

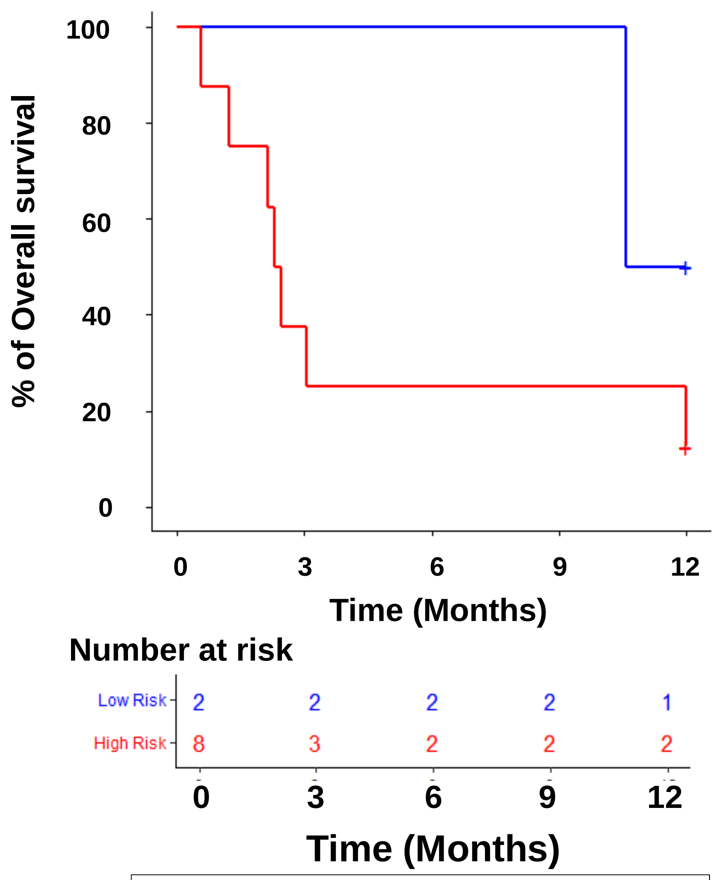


| Variable | Modality | Headcount | HR | CI95% | p-Cox |
|---|---|---|---|---|---|
| Risk groups | | | | | |
| | Low Risk | 2 | 1 | REF | |
| | High Risk | 8 | 3.4 | [0.4- 28] | 0.263 |

#### POP >= 55 & < 60 YO

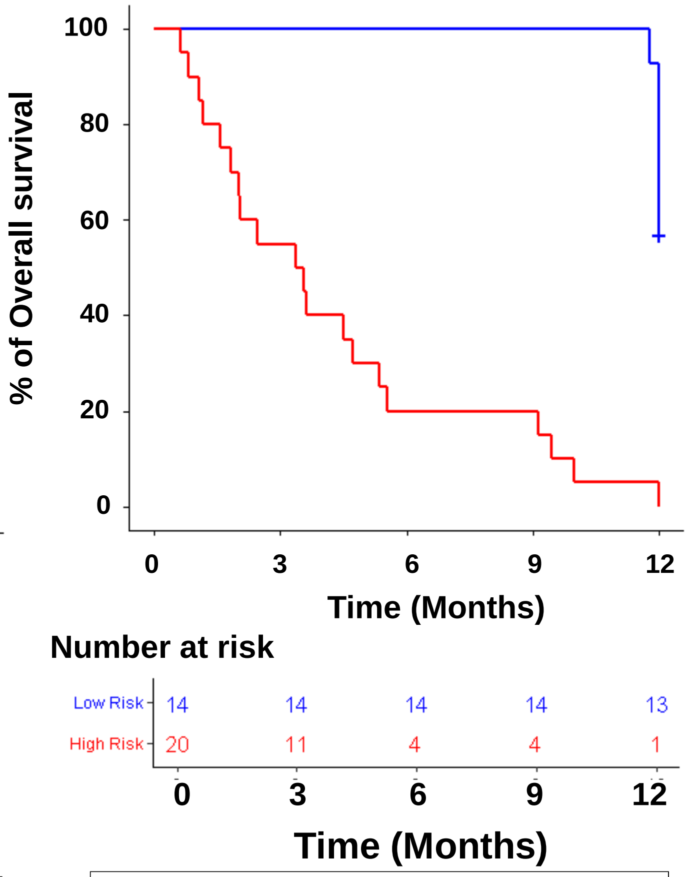


| Variable | Modality | Headcount | HR | CI95% | p-Cox |
|---|---|---|---|---|---|
| Risk groups | | | | | |
| | Low Risk | 14 | 1 | REF | |
| | High Risk | 20 | 15 | [5.2- 44] | < 0.0001 |

#### POP >= 65 & < 75 YO

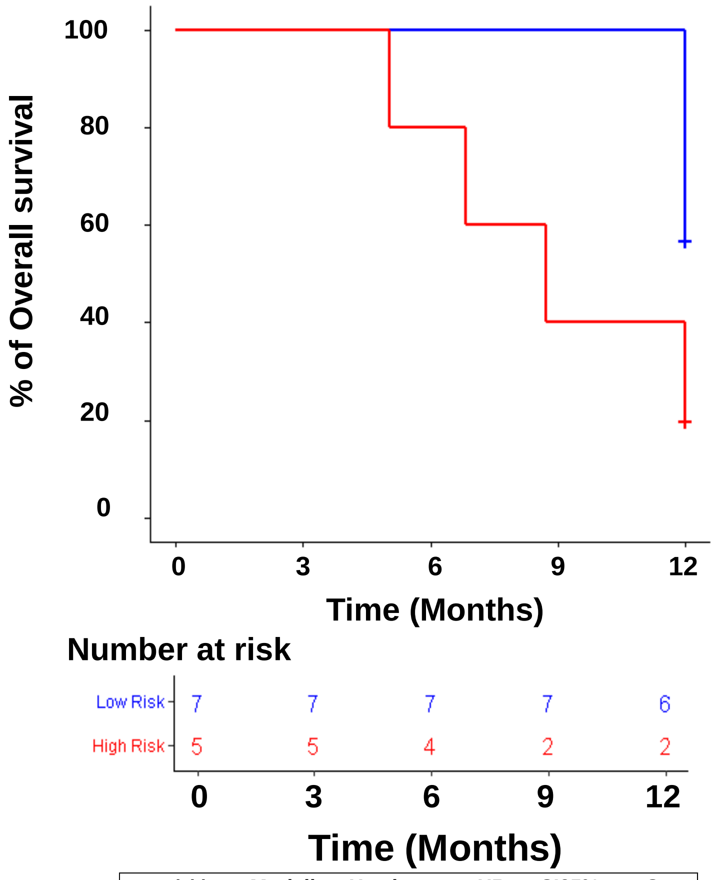


| Variable | Modality | Headcount | HR | CI95% | p-Cox |
|---|---|---|---|---|---|
| Risk groups | | | | | |
| | Low Risk | 7 | 1 | REF | |
| | High Risk | 5 | 3.4 | [0.75- 16] | 0.113 |

**Supplementary Fig. 12: Pace-Locos T, *et al*.**

| Variable | Modality | Number | Frequency |
|---|---|---|---|
| NUMBER OF PATIENTS | | 449 | |
| FURHMAN GRADE | G1 | 14 | 3.2 |
| | G2 | 219 | 49.7 |
| | G3 | 172 | 39 |
| | G4 | 36 | 8.2 |
| SEX | Female | 163 | 36.3 |
| | Male | 286 | 63.7 |
| TUMOR SIZE | T1 | 21 | 4.7 |
| | T1a | 140 | 31.2 |
| | T1b | 108 | 24.1 |
| | T2 | 47 | 10.5 |
| | T2a | 9 | 2 |
| | T2b | 2 | 0.4 |
| | T3 | 3 | 0.7 |
| | T3a | 80 | 17.8 |
| | T3b | 37 | 8.2 |
| | T3c | 2 | 0.4 |
| NODE INVASION | N0 | 201 | 94.8 |
| | N1 | 11 | 5.2 |
| TUMOR STAGE | Stage I | 267 | 59.7 |
| | Stage II | 57 | 12.8 |
| | Stage III | 123 | 27.5 |
| METASTATIC STATUS | NON META | 449 | 100 |
| AGE | mean(sd)<br>median [min-max] | 60.7(12.49) 61[26-90] | |
| OS_STATUS | 0 | 342 | 76.2 |
| | 1 | 107 | 23.8 |
| DFS_STATUS | 0 | 304 | 80.2 |
| | 1 | 75 | 19.8 |
| CLASSE AGE | < 50 y.o | 95 | 21.2 |
| | >= 50 & < 60 y.o | 115 | 25.6 |
| | >= 60 & < 75 y.o | 166 | 37 |
| | >= 75 y.o | 73 | 16.3 |

**Supplementary Table 1: Pace-Locos T, *et al*.**

| Variable | Modality | Headcount | HR | CI95% | p-Cox |
|---|---|---|---|---|---|
| TUMOR STAGE | | | | | |
| | Stage I | 172 | 1 | REF. | - |
| | Stage II | 37 | 1.5 | [ 0.58 - 3.6 ] | 0.424 |
| | Stage III | 85 | 4 | [ 2.3 - 7 ] | <0.0001**** |
| TUMOR PT | | | | | |
| | T1&T2 | 211 | 1 | REF. | - |
| | T3 | 85 | 3.6 | [ 2.1 - 6 ] | <0.0001**** |
| FUHRMAN GRADE | | | | | |
| | G1&G2 | 157 | 1 | REF. | - |
| | G3&G4 | 133 | 2.2 | [ 1.3 - 3.9 ] | 0.00345 ** |

**Supplementary Table 2: Pace-Locos T, *et al*.**

| Variable | Modality | Headcount | HR | CI95% | p-Cox |
|---|---|---|---|---|---|
| TUMOR STAGE | | | | | |
| | Stage I | 149 | 1 | REF. | - |
| | Stage II | 32 | 4 | [ 1.5 - 11 ] | 0.00732 ** |
| | Stage III | 68 | 5.6 | [ 2.4 - 13 ] | <0.0001**** |
| TUMOR PT | | | | | |
| | T1&T2 | 184 | 1 | REF. | - |
| | T3 | 66 | 3.4 | [ 1.7 - 6.7 ] | <0.001*** |
| FUHRMAN GRADE | | | | | |
| | G1&G2 | 139 | 1 | REF. | - |
| | G3&G4 | 106 | 1.6 | [ 0.8 - 3.2 ] | 0.182 |

**Supplementary Table 3: Pace-Locos T, *et al*.**

| Variable | Hazard ratio | HR IC95% low | HR IC95% high | p-value |
|---|---|---|---|---|
| TUMOR_STAGE Stage II | 1.3235 | 0.4937 | 3.5485 | 0.5775 |
| TUMOR_STAGE Stage III | 3.4797 | 1.8955 | 6.3879 | 0.0001 |
| AGE | 1.0426 | 1.0158 | 1.0702 | 0.0017 |
| NARF | 2.1174 | 1.1298 | 3.9682 | 0.0192 |
| UBE2G2 | 3.5283 | 1.5378 | 8.0949 | 0.0029 |
| INTS8 | 2.4770 | 1.3100 | 4.6837 | 0.0053 |
| TMEM167B | 0.4641 | 0.2045 | 1.0535 | 0.0664 |
| ANP32A | 2.6567 | 1.0625 | 6.6431 | 0.0367 |

**Supplementary Table 4: Pace-Locos T, *et al*.**

| Variable | Hazard ratio | HR IC95% low | HR IC95% high | p-value |
|---|---|---|---|---|
| TUMOR_STAGE Stage II | 3.4917 | 1.2331 | 9.8874 | 0.0185 |
| TUMOR_STAGE Stage III | 3.6207 | 1.5125 | 8.6676 | 0.0039 |
| MTAP | 0.3523 | 0.1300 | 0.9548 | 0.0403 |
| C1orf6 | 2.6439 | 0.9995 | 6.9939 | 0.0501 |
| PCDHGC3 | 1.6290 | 0.9097 | 2.9172 | 0.1007 |
| CIAO1 | 5.9043 | 1.3259 | 26.2923 | 0.0198 |
| INTS8 | 3.0408 | 1.3659 | 6.7697 | 0.0065 |

**Supplementary Table 5: Pace-Locos T, *et al*.**

| Variable | Total (%) | Frequency (%) |
|---|---|---|
| Total | 84 | |
| GRADE | | |
| G2 | 10 | 11.9 |
| G3 | 34 | 40.5 |
| G4 | 40 | 47.6 |
| SEX_x | | |
| Female | 25 | 29.8 |
| Male | 59 | 70.2 |
| AJCC_TUMOR_PATHOLOGIC_PT | | |
| T1a | 1 | 1.2 |
| T1b | 3 | 3.6 |
| T2 | 8 | 9.5 |
| T2a | 1 | 1.2 |
| T2b | 2 | 2.4 |
| T3 | 2 | 2.4 |
| T3a | 41 | 48.8 |
| T3b | 15 | 17.9 |
| T4 | 11 | 13.1 |
| META | | |
| META | 84 | 100 |
| AGE | | |
| mean(sd) median [min-max] | 60.25(10.14) 60[33-84] | |
| OS_STATUS | | |
| 0 | 16 | 19 |
| 1 | 68 | 81 |
| AGE_R | | |
| < 50 y.o | 12 | 14.3 |
| >= 50 & < 65 y.o | 23 | 27.4 |
| | 43 | 51.2 |
| >= 75 y.o | 6 | 7.1 |
| AJCC_TUMOR_PATHOLOGIC_PT_R | | |
| T1&T2 | 15 | 17.9 |
| T3 | 58 | 69 |
| T4 | 11 | 13.1 |
| GRADE_R | | |
| G1&G2 | 10 | 11.9 |
| G3&G4 | 74 | 88.1 |

**Supplementary Table 6: Pace-Locos T, *et al*.**
